# The effectiveness of telemedicine in the prevention of type 2 diabetes: a systematic review and meta-analysis of interventions

**DOI:** 10.1101/2024.04.30.24306650

**Authors:** Laura Suhlrie, Raga Ayyagari, Camille Mba, Kjell Olsson, Harold Torres-Aparcana, Steven James, Elpida Vounzoulaki, Daniel B. Ibsen

## Abstract

**Objective:** To evaluate the effectiveness of telemedicine-delivered diet and/or exercise interventions to prevent type 2 diabetes (T2D) in people at risk.

**Methods:** Embase (via Ovid), Medline (via Ovid), Web of Science, CINAHL, Scopus and SciELO were searched from January 2010-December 2020 for intervention studies using a diet and/or exercise intervention delivered through telemedicine for T2D prevention in people at risk. Parallel randomised controlled trials were meta-analyzed and other intervention designs narratively synthesised.

**Results:** We identified 11,645 studies via database searches, of which 226 were full text screened, and 52 interventions included; 32 were included in the meta-analysis and 20 in the narrative synthesis. Telemedicine interventions reduced body weight (mean difference (MD): -1.66 kg, 95% confidence interval (CI) -2.48,-0.90, I^2^=81%, n_studies_=17), body mass index (MD -0.71 kg/m^2^, 95% CI -1.06,-0.37, I^2^=70%, n_studies_=11), waist circumference (MD -2.82 cm, 95% CI -5.16,-2.35, I^2^=84%, n_studies_=7) and HbA1c (MD -0.07%, 95% CI -0.14,0.00, I^2^=71%, n_studies_=11). No significant effects were found for other clinical outcomes. The narrative synthesis supported the results.

**Conclusions:** Our study highlights the potential for telemedicine-delivered interventions in preventing T2D in people at risk.

## INTRODUCTION

Diabetes is the fastest-growing chronic disease world-wide, and is universally acknowledged as posing a significant health, economic, and social burden in all populations. Globally, around 537 million adults aged 20-79 years were estimated to be living with the disease in 2021, with more than 80% of people with diabetes living in low- and middle-income countries (1). Recent projections estimated an increase in global prevalence from 10.5% in 2021 to 12.2% in 2045 (1). Type 2 diabetes (T2D) accounts for approximately 90% of all cases (1). With many health systems currently struggling to meet the demands of diabetes and its sequelae, especially in low- and middle-income countries, it is vital that actions are taken to prevent these projections from becoming a reality.

Weight loss and lifestyle modifications, including a healthy diet and exercise, are cornerstones in T2D prevention (2–4). The American Diabetes Association recommends an intensive behavioural lifestyle intervention program, modelled on the Diabetes Prevention Program (DPP), to achieve and maintain a 7% loss of initial body weight and to increase moderate-intensity physical activity to at least 150 minutes per week (5). The adoption of individual- and group-based in-person interventions to assist with lifestyle modifications is most common in primary care settings, and have been shown to be effective (6). However, common bottlenecks of the success of diabetes prevention programmes include lack of professionally trained staff, limited financial resources, and time constraints, which lead to high absenteeism among participants (7). Similarly, a lack of follow-up in healthcare settings has also been identified (7).

Telemedicine is defined as the “use of information and communication technologies (ICT) for health service delivery” and has been suggested as a viable support tool for primary care physicians for the follow-up of patients with T2D (8, 9). Historically, telemedicine was primarily used to provide care for patients in rural and remote geographical locations (10). In recent years, telemedicine interventions have evolved, and now involve technology such as smartphones, laptop or desk computers to enable communication through voice messages, text and email messages, or live audio or video streaming. During the COVID-19 pandemic, the value of telemedicine was further highlighted, enabling healthcare providers to care for patients remotely (11).

A meta-analysis described DPP-based lifestyle interventions delivered via eHealth and found a mean weight loss of -3.98% (95% confidence interval: -4.49%,-3.46%) from baseline to follow-up of up to 15 months (12), highlighting the effectiveness of telemedicine in supporting weight loss. However, when considering T2D prevention, the effectiveness of other telemedicine interventions, e.g., including different apps and monitoring, to support lifestyle changes and improving multiple health outcomes remains unclear. In addition, to our knowledge there is no evidence on the effectiveness of telemedicine interventions for specific high-risk groups, e.g., people with overweight/obesity, metabolic syndrome or prediabetes. Our systematic review and meta-analysis sought to determine the effectiveness of telemedicine interventions relevant to primary care settings in the prevention of T2D, in people at high risk. We also examined differences in effectiveness among subgroups, including type of control group, degree of behavioural support, type of lifestyle intervention, criteria for being at risk of T2D, risk of bias, and duration of intervention. The protocol of this systematic review and meta-analysis was registered in PROSPERO (CRD42020210829).

## METHODS

A quantitative systematic review was conducted using processes adapted from established review methods set out by the Centre for Reviews and Dissemination (13). Standards derived from the Preferred Reporting Items for Systematic Reviews and Meta-Analysis (PRISMA) were applied (14).

### Literature search methods

Embase (via Ovid), Medline (via Ovid), Web of Science, CINAHL, Scopus and SciELO were systematically searched independently by five authors (EV, KO, CM, HT and SJ). All articles published between January 2010-December 2020 were retrieved. The rationale behind including only articles published after 2010 is that broadly available telemedicine technologies emerged around this time period (15, 16). The MESH headings ‘glucose Intolerance [mh]’, ‘Telemedicine [mh]‘, ‘Mobile Applications [mh]‘ and ‘Cell Phone [mh]‘, ‘Text Messaging [mh]‘, ‘Internet-Based Intervention [mh]‘, ‘Internet of Things [mh]‘, ‘Internet [mh]‘, ‘Primary Prevention [mh]‘, ‘Life Style [mh]‘, ‘Healthy Lifestyle [mh]‘, ‘Risk Reduction Behavior [mh]‘, ‘Sedentary Behavior [mh]‘, ‘Diet [mh] Exercise [mh]‘, ‘habits [mh]‘, ‘Primary Health Care [mh], ‘Care Nursing [mh]‘, ‘Nutritionists [mh]‘, ‘Physical Therapists [mh], and ‘Nurses [mh]‘ were used. Full details of the search strategy for Medline (via OVID) that was then adapted for the other database searches are provided in **Supplemental Table 1**. The search strategy used was initially designed by five authors (EV, KO, CM, HL and SJ) and revised by a librarian with expertise in conducting systematic reviews and meta-analyses. Articles identified from database searches were exported to EndNote (Clarivate Analytics) where duplicates were removed. Duplicates were further identified and removed after import into Covidence. Titles and abstracts were then independently screened by three pairs of researchers (RA, CM, KO, HT, SJ and DBI). Full-text articles were obtained if the abstracts were considered eligible by at least two reviewers. Each full text was then independently screened by two reviewers for final inclusion; if an agreement could not be reached, then conflicts at each stage were resolved by a third independent researcher. Reference lists of relevant studies were hand-searched to identify additional potentially eligible studies for inclusion. We corresponded with the study authors to request missing data required to conduct the meta-analysis.

### Eligibility criteria

Inclusion criteria comprised adult populations at high risk of developing T2D (diagnosis of prediabetes confirmed by clinical parameters or screening tools, metabolic syndrome, or overweight/obesity). Interventions assessed involved any technologically assisted primary prevention strategies, including video conferencing, text messages, e-mail, internet, smartphone applications, phone calls. No restrictions were made regarding the control group (if available). Any clinically relevant outcomes related to metabolic syndrome were considered for the meta-analysis. The primary outcome was change in body weight. For the meta-analysis, only randomised controlled trials (RCTs) were included. We included non-randomized designs in the narrative review. No limitations were made regarding the language of the study and country where the intervention took place as long as the intervention setting was primary care or was relevant to primary care (17) (**Supplemental Table 2**).

Studies including populations aged less than 18 years old, adults with known diabetes, and adults not at high risk of developing T2D were excluded. Furthermore, interventions not delivered via telemedicine were excluded. Intervention types consisting of other means than primary prevention or targeting other types of diabetes than T2D (i.e. type 1 diabetes (T1D), gestational diabetes, atypical diabetes) were not considered in this review. Any observational studies, literature reviews, commentaries, opinion pieces, narrative overviews, and qualitative studies were also excluded, as were studies that did not take place or were not relevant to the primary care sector (e.g. studies including hospitalised patient populations).

### Risk of bias assessment

Risk of bias assessment was undertaken by three authors (SJ, KO and LS) using the Cochrane risk of bias tool (18) (**Supplemental Table 3**). To ensure reliability in data extraction and quality appraisal, a sample of papers included in the review were independently appraised and data extraction was compared by three additional authors (DBI, RA and LS). Agreement was reached for all papers.

### Data extraction and synthesis

Study information was extracted using a standardised form created in Covidence and data for meta-analyses were compiled in an Excel spreadsheet. Data extracted included: Study ID, outcome, estimate description, estimate value, confidence intervals, standard error, p-value, criteria for high-risk of T2D, intervention setting, telemedicine strategy used, medium of communication used, lifestyle intervention focus, intervention duration, degree of behavioural support, type of control group, number of participants, mean baseline age, percentage female, ethnicity. Categorizations informed by previous research were performed to generate subgroups (19), including criteria for high-risk of T2D (overweight/obese, prediabetes, prediabetes and overweight/obese, metabolic syndrome, other), intervention setting (home, hospital, pharmacy, other), telemedicine strategy (taxonomy as classified by Lee et al. (19): teleconsultation, tele-education, telecase-management, telemonitoring, telementoring, tele-education & telecase-management, tele-education and telemonitoring, tele-management & telemonitoring, tele-management and teleconsultation), medium of communication used (App, internet, short messages, telephone), lifestyle intervention focus (diet, diet and exercise, diet, exercise and medication, and exercise alone), intervention duration in months, degree of behavioural support (no support, supported by face-to-face contact, supported by remote and face-to-face contact, and supported by remote contact), type of control (usual care, wait list control, usual care and minimal telemedicine only).

### Statistical analysis

Identified interventions were grouped according to the design of the studies as follows: parallel-group RCTs, other types of RCTs (e.g. cross-over or cluster RCTs) and non-randomized and non-controlled interventions. For consistency, only parallel-group RCTs were meta-analysed. Other study designs were used as supporting evidence and included in the narrative synthesis.

Random-effects meta-analytic models were fitted using Bayesian methods, as these have been suggested to be more stable than frequentist methods (i.e. DerSimonian and Laird), particularly when notable between-study heterogeneity is present and when the number of studies is small (20). All models were fitted using R version 4.3.1, using the ‘bayesmeta’ (ref), ‘metaviz’, and ‘meta’ packages (21). The prior for the mean effect was set conservatively as 0 and the standard deviation of the prior was set as the maximum observed standard deviation among the included studies for each outcome (22).

Heterogeneity was quantified using the proportion of variability that is due to between-study heterogeneity (tau) and within-study variability (Higgins’ I^2^). A 95% confidence interval (CI) was estimated for tau, rather than a point estimate, as these are known to be difficult to estimate unless the number of studies is large. We used the half-Caucy distribution for the heterogeneity (tau), set at 1, which is considered “fairly high” (22).

Subgroup meta-analyses were performed using the standard inverse variance methods. -Subgroups included: 1) comparison groups, 2) degree of behavioural support, 3) type of intervention, 4) type of population, 5) risk of bias, and 6) duration of intervention. The type of comparison group was investigated, as this is important for the interpretation of results and relevance of the interventions. The degree of behavioural support and type of population was investigated as each intervention and population subtype has different implications in primary care settings. Because we used a broad definition of telemedicine interventions, the interventions were likely to vary substantially and use different types of technology. Thus, we investigated the subtypes of telemedicine interventions. Risk of bias was investigated to assess how study bias may impact the estimates. Lastly, the duration of the intervention was investigated to assess long-term adherence.

In sensitivity analyses, we used different prior distributions for the heterogeneity, using either the half-Cauchy or half-normal distribution with different scales. Publication bias was evaluated using funnel plots, Egger’s test and the trim-and-fill-method (23, 24).

## RESULTS

### Search results

A total of 15,158 records were retrieved in Endnote with 1660 duplicates identified; 13,498 studies were uploaded into Covidence and further 1853 duplicates identified. In total, 11,645 studies were identified via electronic database searches Embase (via Ovid), Medline (via Ovid), Web of Science, CINAHL, Scopus and SciELO, with 226 included for full text screening (**Figure 1**). Full text screening resulted in 52 eligible studies being includedt, of which 32 parallel RCTs were eligible for inclusion in the meta-analyses and 20 studies in the narrative synthesis (four cluster-randomised controlled trials, seven non-randomized controlled trials and nine non-controlled trials).

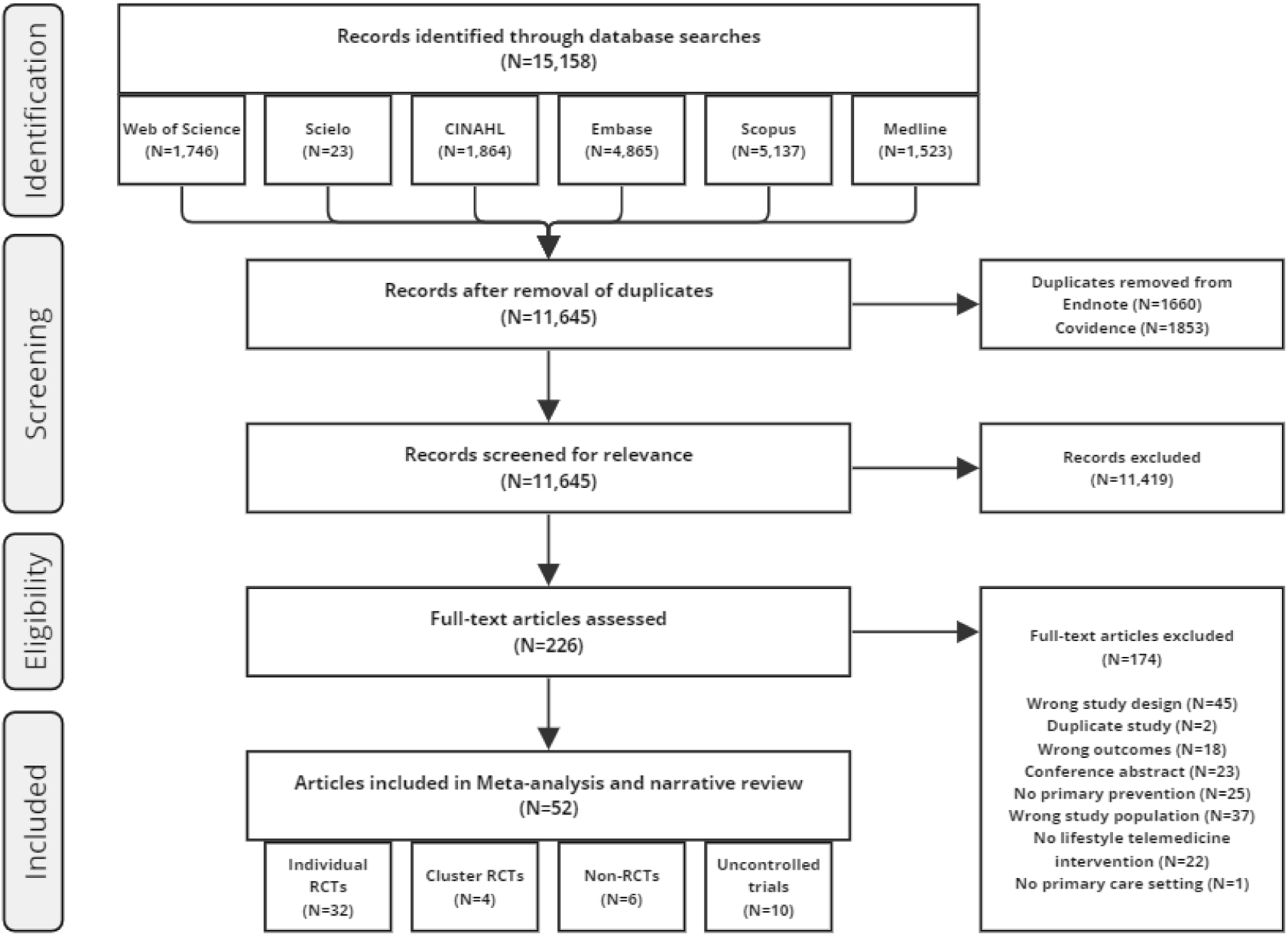

### Study characteristics

Of the 32 RCTs included in the meta-analysis (**Table 1**), most (15/32) included participants based on being overweight or obese, performed the intervention in the home setting (13/32) in a Western country (26/32), focused on diet and exercise (24/32), included tele-monitoring and/or tele-education (30/32), were supported by remote contact (20/32), and compared the telemedicine intervention with usual care (15/32).

**Table 1.** Characteristics of individually randomised intervention studies included in the meta-analysis investigating effectiveness of telemedicine intervention on prevention of type 2 diabetes.

| Study, year (ref) | Criteria for high-risk | Setting | Region | Intervention focus | Telemedicine | Support | Control group | Intervention duration | N participants | Risk of bias assessment |
| --- | --- | --- | --- | --- | --- | --- | --- | --- | --- | --- |
| Aguiar 2016 (51) | Overweight/obesity | Home | Australia | Diet and exercise | Tele-education, Telemonitoring | No support | Wait list control | 6 | 101 | Low |
| Allman-Farinelli 2016 (52) | Overweight/obesity | Home | Australia | Diet and exercise | Tele-education, Telemonitoring | Supported by remote contact | Usual care + minimal telemedicine only | 3 | 250 | High |
| Apinaniz 2019 (53) | Overweight/obesity | Combination | Europe | Diet and exercise | Tele-education | Supported by remote contact | Intervention without telemedicine | 6 | 110 | Some |
| Bender 2018 (54) | Prediabetes & overweight | Combination | North America | Diet and exercise | Tele-education, Telemonitoring, Telementoring | Supported by remote | Wait list control | 3 | 67 | Some |
| Block 2015 (55) | Prediabetes & overweight | Home | North America | Diet and exercise | Tele-education, Telemonitoring | No support | Wait list control | 6 | 339 | Low |
| Bosak 2010 (56) | Metabolic syndrome | Combination | North America | Exercise | Tele-education, Telemonitoring | Supported by remote contact | Usual care | 1.5 | 22 | High |
| Carnie 2013<br>(57) | Overweight/obesity | Pharmacy | North America | Diet and exercise | Tele-education | Supported by remote and face-to-face contact | Usual care + minimal telemedicine only | 6 | 199 | Low |
| Chen 2020<br>(58) | Prediabetes | Combination | Asia | Diet | Combination | Supported by remote and face-to-face contact | Usual care | 3 | 138 | Some |
| Cho 2020<br>(59) | Metabolic syndrome | Home | Asia | Diet and exercise | Telementoring, Telemonitoring | Supported by remote contact | Usual care | 6 | 129 | Low |
| Cicolini 2014<br>(60) | Hypertension | Clinic | Europe | Diet, exercise and medication | Tele-education | Supported by remote contact | Usual care | 6 | 203 | Low |
| Fischer 2015<br>(61) | Prediabetes & overweight | Combination | North America | Diet and exercise | Tele-education | Supported by remote contact | Usual care | 12 | 163 | High |
| Fukuoka 2015<br>(62) | Prediabetes & overweight | Combination | North America | Diet and exercise | Tele-education, Telemonitoring | Supported by face-to-face contact | Usual care | 5 | 61 | High |
| Green 2014<br>(63) | Overweight/obesity | Clinic | North America | Diet | Tele-education, Telemonitoring | Supported by remote contact | Usual care | 6 | 101 | Some |
| Hebden 2014 (64) | Overweight/obesity | Home | Australia | Diet and exercise | Tele-education, Telemonitoring | No support | Usual care | 3 | 51 | Some |
| Hurkmans 2018 (65) | Overweight/obesity | Home | Europe | Diet and exercise | Other | Supported by face-to-face contact | Wait list control | 3 | 102 | High |
| Jahangiry 2015 (66) | Metabolic syndrome | Home | Asia | Diet and exercise | Tele-education, Telemonitoring | Supported by remote contact | Wait list control | 6 | 160 | High |
| Johnson 2019 (67) | Overweight/obesity | Home | North America | Diet and exercise | Telemonitoring, Telementoring | Supported by remote contact | Usual care + minimal telemedicine only | 3 | 30 | Low |
| Kempf 2018 (68) | Overweight/obesity | Other | Europe | Diet and exercise | Tele-education, Telemonitoring, Telementoring | Supported by remote contact | Usual care | 3 | 180 | Low |
| Lison 2020 (69) | Overweight/obesity | Hospital | Europe | Diet and exercise | Tele-education, Telemonitoring | No support | Wait list control | 3 | 105 | Low |
| Ma 2013 (70) | Prediabetes & overweight | Clinic | North America | Diet and exercise | Tele-education, Telemonitoring | Supported by remote contact | Usual care | 15 | 241 | Low |
| McLeod 2020 (71) | Prediabetes | Combination | Australia | Overall behaviour | Tele-education, Telemonitoring, Telementoring | Supported by remote contact | Usual care | 12 | 225 | Low |
| Patel 2019<br>(72) | Overweight/obesity | Home | North America | Diet | Tele-education, Telemonitoring | Supported by remote contact | Intervention + minimal telemedicine only | 6 | 105 | High |
| Pérez Ewert 2016<br>(73) | Prediabetes & overweight | Home | South America | Diet and exercise | Tele-education, Telemonitoring | Supported by remote contact | Usual care | 9 | 70 | Low |
| Peyer 2017<br>(74) | Overweight/obesity | Combination | North America | Diet and exercise | Telemonitoring | Supported by face-to-face contact | Intervention without telemedicine | 2 | 89 | Some |
| Rogers 2016<br>(75) | Overweight/obesity | Combination | North America | Diet and exercise | Tele-education, Telemonitoring | Supported by remote contact | Intervention without telemedicine | 6 | 39 | High |
| Silina 2017 | Overweight/obesity | Home | Europe | Diet and exercise | Tele-education | No support | Intervention without telemedicine | 12 | 129 | Some |
| Staite 2020<br>(76) | Prediabetes & overweight | Home | Europe | Diet and exercise | Tele-education, Telemonitoring | No support | Usual care + minimal telemedicine only | 12 | 200 | High |
| Tanaka 2018<br>(77) | Prediabetes & overweight | Home | Asia | Diet | Tele-management, Telemonitoring | Supported by remote contact | Usual care | 3 | 112 | Low |
| Tarraga Marcos 2017<br>(78) | Overweight/obesity | Clinic | Europe | Diet | Tele-education, Telemonitoring | Supported by face-to-face contact | Usual care | 12 | 120 | Some |
| Toro-Ramos<br>2020<br>(79) | Prediabetes | Home | North<br>America | Diet and<br>exercise | Tele-education,<br>Teleconsultation,<br>Telemonitoring | Supported<br>by remote<br>contact | Intervention<br>without<br>telemedicine | 12 | 202 | High |
| Weinstock<br>2013<br>(80) | Metabolic<br>syndrome | Home | North<br>America | Diet and<br>exercise | Tele-education, Telecase-management | Supported<br>by remote<br>contact | Usual care +<br>minimal<br>telemedicine<br>only | 24 | 257 | High |
| Wu 2011<br>(81) | Overweight/obesity<br>& other risk factors | Home | Asia | Diet and<br>Exercise | Tele-education,<br>Teleconsultation,<br>Telemonitoring | Supported<br>by remote<br>contact | Usual care | 9 | 135 | High |

In total, 12 of the 32 studies were judged to be at high risk of bias, 8 to have some risk of bias and 12 to have low risk of bias (**Supplemental Table 3**). The most common reason for being at high risk of bias was incomplete outcome data.

For the narrative synthesis, we identified 4 cluster-randomised studies, 6 non-randomized studies, and 10 non-controlled interventions. Of those 20 studies (**Table 2**), the majority included populations with prediabetes (6/20), followed by populations with prediabetes and overweight/obesity (4/20) and populations with overweight/obesity only (4/20). The remaining studies included populations with other risk factors such as metabolic syndrome (3/20). The intervention setting comprised home (5/20), community (5/20), clinic (4/20), workplace (2/20) and a combination of different settings (4/20). Ten studies were conducted in Asia, seven studies in North America and three studies in Europe. Like the RCTs included in the meta-analysis, most studies in the narrative analysis emphasized diet and exercise interventions. Most focused on both diet and exercise (16/20), and some on diet only (3/20) or exercise only (1/20). One study involved medication management.

**Table 2.** Characteristics of cluster-randomised, non-randomised and non-controlled intervention studies excluded from the meta-analysis investigating effectiveness of telemedicine intervention on prevention of type 2 diabetes.

| Study, year (ref) | Design | Criteria for high-risk | Setting | Region | Intervention focus | Telemedicine | Support | Control group | Duration, months | N | Results summary |
| --- | --- | --- | --- | --- | --- | --- | --- | --- | --- | --- | --- |
| Chee 2014 (27) | Cluster randomised parallel group design | Metabolic syndrome | Home | Asia | Exercise | Tele-education | Supported by face-to-face contact | Intervention without telemedicine | 4 | 147 | Within-group analyses revealed that the intervention group significantly increased the step number per day with a stronger increase between baseline and two months compared to between two months follow up and four months, |
| Davies 2016 (25) | Cluster randomised parallel group design | Prediabetes | Clinic | Europe | Diet and exercise | Tele-education | Supported by remote and face-to-face contact | Usual care | 36 | 880 | A 6-hour intervention followed by yearly 3-hour refreshers showed a non-significant 26% reduced risk of developing T2DM and statistically significant improvements in HbA1c (-0.06%, CI[-0.11, -0.01]), LDL cholesterol (-0.08 mmol/mol CI[-0.15, -0.01]), sedentary time (-26.29 min, CI[-45.26, -7.32]) and step count (498.15, CI[162.10, 834.20]). |
| Rusali 2018 (26) | Cluster randomised parallel group design | Overweight/obesity | Workplace | Asia | Diet and exercise | Tele-education | No support | Usual care group and face-to-face group | 4 | 108 | The online intervention group showed lower reductions in body weight (-1.12 kg) compared to the face-to-face intervention group (-5.80 kg) and control group (-1.82 kg) and similar trends were observed in BMI (-2.6 kg/m<sup>2</sup> in face-to-face group vs. -0.4 kg/m<sup>2</sup> in telemedicine group vs. -1.1 kg/m<sup>2</sup> in control group) (p-value interaction effect <0.01). |
| Sakane 2019 (28) | Cluster randomised parallel group design | Prediabetes | Community | Asia | Diet and exercise | Tele-education, Telemonitoring | Supported by remote contact | Usual care + minimal telemedicine only | 12 | 1597 | It was observed that 8.0% of the participants in the intervention group and 12.0% in the control group developed Metabolic Syndrome. In the overall analysis, the hazard ratio for the development of metabolic syndrome in the intervention group was insignificant, however, when focusing on individuals who were overweight or obese the hazard ratio was significantly reduced to 0.63 (95% CI: 0.41–0.95, P=0.029). |
| Ciemins 2018 (29) | Non randomised controlled trial | Prediabetes & overweight | Combination | North America | Diet and exercise | Tele-education | Supported by remote and face-to-face contact | Intervention without telemedicine | 4 | 667 | The modified group DPP showed overall improvements in reaching 5% and 7% weight loss. There were no significant differences between the face-to-face and telehealth groups (33.5% and 34.6% for 7% weight loss; 49.6% and 46.6% for 5% weight loss). |
| Kim 2019 (32) | Non randomised controlled trial | Metabolic syndrome | Combination | Asia | Diet and exercise | Tele-education, Telemonitoring | Supported by remote and face-to-face contact | Intervention without telemedicine | 6 | 1117 | Preference for low-sodium diet, reading nutritional facts, having breakfast, and performing moderate physical activity significantly increased in the mobile health intervention group. |
| Laitinen 2010 (34) | Non randomised controlled trial | Other | Combination | Europe | Diet and exercise | Tele-education | No support | Intervention without telemedicine | 21 | 74 | The outcomes of the intervention showed mostly no significant differences between both study groups. Waist circumference decreased significantly between baseline and 6 months of counselling and was significantly lower at 21 months than at baseline in the telemedicine intervention group. |
| Moin 2018 (30) | Non randomised controlled trial | Prediabetes & overweight/obesity | Community | North America | Diet and exercise | Tele-education, Telemonitoring | Supported by remote contact | Intervention without telemedicine | 12 | 268 | The online DPP showed significant reductions in weight at 6 and 12 months with -4.7 kg [-6.5 kg, -2.8] and -4.0 kg [-4.9 kg, -3.0 kg] respectively, and no significant difference compared with in-person DPP. |
| Toro-Ramos 2017 (31) | Non randomised controlled trial | Overweight/obesity | Home | Asia | Diet and exercise | Tele-education, Telemonitoring | Supported by remote contact | Intervention without weight loss curriculum | 3.8 | 159 | The study showed that participants achieved a significant reduction in body weight of 7.5%, in percent body fat (ranging from -6.0% to -5.4%) and visceral fat (decreasing from -3.4 kg to -2.7 kg) at the end of a 15-week program as well sustained effects at 52-week follow-up (5.2% weight loss). |
| Vadheim 2010 (33) | Non randomised controlled trial | Overweight/obesity & other diabetes risk factors | Community | North America | Diet and exercise | Tele-education, Telemonitoring | Supported by remote contact | Intervention without telemedicine | 4 | 27 | On average, both study arms increased physical activity levels and reduced weight by at least 7% with no significant differences between the groups. |
| Alwashmi 2019 (35) | Uncontrolled trial | Prediabetes | Combination | North America | Diet and exercise | Tele-education, Telemonitoring | Supported by remote contact | No control group | 4 | 273 | This study investigated the impact of participation in the Transform DPP program and found significant reductions in weight (-13.3 lbs or -6.5%), BMI (-1.9 kg/m<sup>2</sup>), increased exercise frequency (1.7 days per week), and reduced work absenteeism. |
| An 2019 (36) | Uncontrolled trial | Overweight/obesity | Home | Asia | Diet | Telemonitoring | Supported by remote contact | No control group | 2.5 | 40 | The outcomes of the intervention revealed significant reductions in body weight (-0.6 kg), BMI (-0.2 kg/m<sup>2</sup>), waist circumference (-1.4 cm), and body fat mass (-0.7 kg) among all the participants as well as significant improvements in anthropometric parameters, glycemic control, and lipid profile in the high-adherence-group. |
| Cameron Sepah 2017 (37) | Uncontrolled trial | Prediabetes & overweight/obesity | Home | North America | Diet and exercise | Tele-education, Telemonitoring, Telementoring | Supported by remote contact | No control group | 36 | 220 | Participants who completed four or more lessons and those who completed nine or more lessons of the digital DPP achieved sustained weight loss (-3.0% and -2.9%, respectively) and experienced a notable reduction in HbA1c levels (-0.31 and -0.33, respectively). |
| Chen 2014 (38) | Uncontrolled trial | Prediabetes | Clinic | Asia | Diet and exercise | Telecase-management | Supported by remote and face-to-face contact | No control group | 6 | 2219 | Patients with prediabetes taking part in a Smart Web Aid for Preventing Type 2 Diabetes showed improvements from baseline to 6-months follow-up in vegetable intake, calorie intake, leisure-time exercises, body weight, and body mass index as well as improvement in self-efficacy for modifying diet, increasing physical activities, engaging relatives, and knowledge about diabetes and risks of an imbalanced diet and inadequate physical activity. |
| Choi 2014 (39) | Uncontrolled trial | Other | Community | Asia | Diet | Tele-education | Supported by remote contact | No control group | 4 | 29 | Intervention outcomes showed significant decreases in body weight, waist circumference, and BMI among female and male participants. The frequency of certain dietary habits such as snacking, eating out, and dining with others decreased, while the frequency of consuming foods like whole grains, seaweed, fruit, and low-fat milk increased. |
| Kim 2019 (40) | Uncontrolled trial | Prediabetes | Community | North America | Diet and exercise and medication | Tele-education, Telemonitoring | Supported by remote and face-to-face contact | No control group | 6 | 247 | The feasibility study showed significant improvements of clinical outcomes (e.g., HbA1c, weight, depression, and blood pressure, cholesterol) and no one with prediabetes progressed to diabetes. |
| Michaelides 2016 (41) | Uncontrolled trial | Prediabetes & overweight/obesity | Home | North America | Diet and exercise | Tele-education, Telemonitoring | Supported by face-to-face contact | No control group | 6 | 54 | Results of the virtual DPP showed significant weight reductions at 4 and 6 months ($-5.4 \pm 4.43$ kg and $6.22 \pm 5.00$ kg respectively) follow-up with people who completed the program showing stronger engagement and more weight loss ($-6.0 \pm 4.34$ kg and $-7.01 \pm 4.83$ kg). |
| Noh 2020 (44) | Uncontrolled trial | Prediabetes | Workplace | Asia | Diet and exercise | Tele-education, Telemonitoring | Supported by remote contact | No control group | 3 | 62 | The authors found significantly reduced HbA1c values by $-0.38 \pm 0.65\%$ in all participants and stronger reductions in the participant group with HbA1c above 6.4% at baseline ($-0.78 \pm 0.79\%$). |
| Rossi 2010 (42) | Uncontrolled trial | Overweight/obesity | Clinic | Europe | Diet | Tele-education, Telemonitoring | Supported by remote contact | No control group | 5 | 140 | The mobile phone based intervention resulted in an increased percentage of participants reaching Mediterranean diet targets (14.4% to 69.8%), a mean (95% confidence interval) reduction in body weight by 2.5 (3.2; 1.8) kg, whereas the reduction in waist circumference and BMI were 3.7 (4.6; 2.9) cm, and 1.0 (0.7; 1.2) kg/m<sup>2</sup>. |
| Song 2011 (43) | Uncontrolled trial | Metabolic syndrome | Clinic | Asia | Diet and exercise | Tele-management, Teleconsultation | Supported by remote contact | No control group | 6 | 316 | Significant reductions in blood pressure, weight, abdominal visceral fat, physical age, the extent of walking, waist size and blood sugar ( $p < 0.01$ );<br>body fat and BMI ( $p < 0.05$ );<br>and body fat (triacylglycerol; $p < 0.1$ ) were found after the<br>implementation of a metabolic<br>syndrome care service model. |

Most telemedicine strategies included education and monitoring (10/20) as well as only tele-education (5/20). Furthermore, the intervention groups mostly received remote support (11/20) as well as remote and face to face support (5/20). The intervention duration was 9 months on average (range between 2.5 months and three years) and 432 participants on average (range between 27 and 2219).

### Randomised controlled trials

Using telemedicine in people at risk of T2D reduced body weight (mean difference (MD) -1.66 kg, 95% confidence interval (CI) -2.48, -0.90, I^2^ = 81%, n studies = 17), body mass index (BMI) (MD -0.71 kg/m^2^, 95% CI -1.06, -0.37, I^2^ = 70%, n studies = 11) and waist circumference (MD -2.82 cm, 95% CI -5.16, -2.35, I^2^ = 84%, n studies = 7), though with substantial heterogeneity (**Table 3, Supplemental Figure 1**). In subgroup analyses of body weight changes, the largest weight reductions were found when comparing the telemedicine intervention to wait list controls or intervention without telemedicine, as opposed to usual care or minimal telemedicine (**Table 4**). Telemedicine interventions in people with overweight/obesity showed greater average weight reductions than in people with prediabetes, prediabetes and overweight/obesity, or metabolic syndrome (**Table 4**).

**Table 3.**
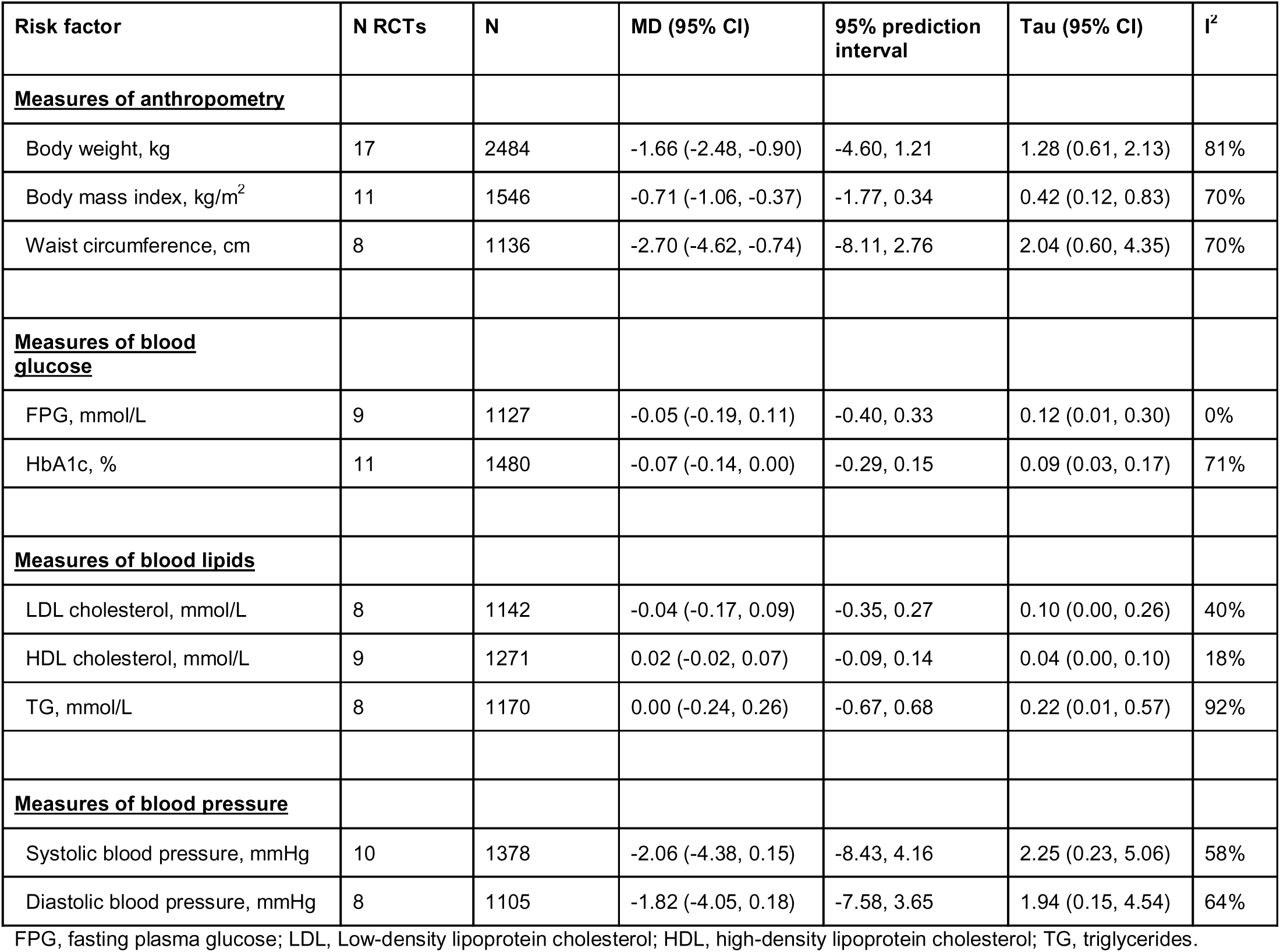
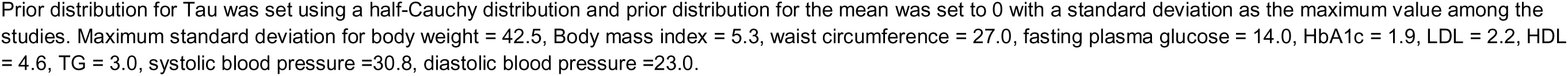
Overview of pooled results from randomised controlled trials investigating telemedicine for prevention of type 2 diabetes.

**Table 4.** Subgroup meta-analysis of randomized controlled trials delivering lifestyle interventions using telemedicine to reduce body weight

| Subgroup | N RCTs | MD (95% CI) | I<sup>2</sup> |
| --- | --- | --- | --- |
| **<u>Overall</u>** | 17 | -1.66 (-2.48, -0.90) | 81% |
| **<u>Type of control group</u>** |  |  |  |
| Intervention without telemedicine | 3 | -2.48 (-4.48, -0.47) | 0% |
| Usual care | 8 | -1.26 (-2.18, -0.34) | 32% |
| Minimal telemedicine | 3 | -1.09 (-2.81, 0.63) | 73% |
| Wait list control | 3 | -2.74 (-4.29, -1.19) | 92% |
| **<u>Degree of behavioral support</u>** |  |  |  |
| No support | 4 | -2.07 (-3.60, -0.54) | 89% |
| Supported by remote contact | 12 | -1.58 (-2.46, -0.70) | 44% |
| Supported by remote and face-to-face contact | 1 | -1.40 (-4.06, 1.26) | - |
| **<u>Type of lifestyle intervention</u>** |  |  |  |
| Overall behaviour | 1 | -0.50 (-3.16, 2.16) | - |
| Diet | 4 | -1.57 (-2.98, -0.17) | 40% |
| Diet and exercise | 12 | -1.84 (-2.71, -0.97) | 73% |
| **<u>Population at high-risk</u>** |  |  |  |
| Overweight/obese | 7 | -2.64 (-3.81, -1.47) | 75% |
| Prediabetes | 3 | -1.19 (-2.59, 0.20) | 24% |
| Prediabetes and overweight/obese | 4 | -0.90 (-2.16, 0.35) | 18% |
| Metabolic syndrome | 3 | -1.57 (-2.99, -0.15) | 55% |
| **<u>Risk of bias</u>** |  |  |  |
| Low | 5 | -1.84 (-3.12, -0.56) | 83% |
| Some | 7 | -1.57 (-2.74, -0.40) | 53% |
| High | 5 | -1.65 (-3.05, -0.25) | 67% |

| <u>Duration of intervention</u> |  |  |  |
| --- | --- | --- | --- |
| Short (<6 months) | 5 | -1.34 (-2.45, -0.19) | 45% |
| Medium (6 to <12 months) | 5 | -2.47 (-3.70, -1.24) | 79% |
| Long (>= 12 months) | 6 | -1.31 (-2.41, -0.20) | 54% |

Interventions between 6 to 12 months also showed slightly larger weight reductions than shorter or longer duration interventions (**Table 4**). No substantial differences were observed with different degrees of behavioural support, type of lifestyle intervention, or risk of bias (**Table 4**). Similar patterns were observed for type of control group and intervention duration for BMI and waist circumference (**Supplemental Tables 4-5**). A greater average reduction in BMI was observed for studies with low risk of bias compared to some or high risk of bias (**Supplemental Table 4**).

There was no evidence of an effect of the telemedicine-delivered intervention on fasting plasma glucose (FPG) compared with different comparison groups (**Table 3, Supplemental Figure 1**). A reduction in HbA1c by 0.07% (95% CI -0.14, 0.00, I^2^ = 71%) was observed among the 11 included studies with available data (**Table 3**). In subgroup analyses, we observed a reduction in FPG and HbA1c among telemedicine interventions supported by remote contact and graded as low risk of bias (**Supplemental Tables 6-7**). The greatest reduction in HbA1c was observed among people with overweight/obesity (MD -0.13%, 95% CI -0.21, -0.05, I^2^ = 28%). A reduction in HbA1c was found in short and medium duration intervention (<12 months) but not among long-term studies (≥12 months).

No statistically significant differences between telemedicine interventions and comparison groups for low-density lipoprotein (LDL) cholesterol, high-density lipoprotein (HDL) cholesterol and triglycerides were found in the overall analysis (**Table 3, Supplemental Figure 1**). Similarly, no statistically significant differences were observed in subgroup analyses (**Supplemental Table 8-10**). The exception was that one study found reduced triglycerides when the telemedicine intervention was supported by remote and face-to-face contact, whereas two studies found increased triglycerides when compared to no support (**Supplemental Table 10**).

No statistically significant difference was observed for systolic or diastolic blood pressure in the overall meta-analysis comparing telemedicine intervention with different comparison groups (**Table 3, Supplemental Figure 1**). Reduced systolic and diastolic blood pressure was found among interventions focusing on both diet and exercise (systolic blood pressure - 1.91 mmHg, 95% CI -3.81, -0.01), I^2^ = 57%), among participants with prediabetes (systolic blood pressure -3.31 mmHg, 95% CI -6.48, -0.17, I^2^ = 63%) and in studies with high risk of bias (**Supplemental Tables 11-12**).

In sensitivity analyses, different prior distributions were set. The overall mean differences were like those estimated in the main analyses (**Supplemental Table 13**). Publication bias was investigated using Egger’s test, trim-and-fill analysis and visually by inspection of funnel plots (**Supplemental Table 14 and Supplemental Figure 2**). Egger’s test indicated publication bias in the analysis of body weight (p = 0.043) and trim-and-fill analysis suggested a lower reduction in body weight (MD -0.95 kg, 95% CI -1.80, -0.11). Trim-and-fill analysis also suggested estimates closer to the null for systolic and diastolic blood pressure (**Supplemental Figure 2**).

### Cluster-randomised controlled, non-controlled and non-randomized interventions

In the cluster-randomised controlled trials, significant improvements in clinical and anthropometric parameters were found in the telemedicine intervention compared to the control groups (25, 26). However, in one study, reductions in weight and BMI were greater in the face-to-face intervention delivery mode compared to a telemedicine intervention without behavioural support (26). Two studies found that participants participating in telemedicine interventions had significantly increased physical activity and less sedentary time compared to controls (25, 27). Further, two studies found that telemedicine interventions improved dietary behaviour (25, 26). One study found a significantly lower risk of developing metabolic syndrome in overweight or obese individuals (28). Davies et al. found significant improvements in illness perception rates, quality of life, and anxiety but no significant changes related to depression, and sleep behaviour (25).

Within the non-randomized controlled trials, mobile health interventions to decrease metabolic risk significantly improved clinical (29–31) and health behaviour (32, 33) primary outcomes. The studies showed improvements in secondary outcomes such as in blood pressure, FPG, triglycerides, and HDL (31, 34), but only one study had statistically significant changes (31). Regarding anthropometric improvements, two studies showed significant improvements of clinically relevant 5% weight loss (29, 31). Although they had smaller effect sizes, two studies showed maintained weight loss effects after one year (30, 31). Improved health behaviours included significant improvements related to nutrition (e.g. preference for low sodium diet, having breakfast) (32) and increased moderate physical activity (32, 33). In three out of five studies, no significant between-group effect differences were found between telehealth intervention groups compared to face-to-face comparison groups (29, 30, 32–34). In one study waist circumference significantly decreased in the face to face group compared to a telehealth intervention but other between-group differences were not significant (34). In another study health behaviour improved significantly in a telehealth intervention group compared to a face-to-face group (32).

Within the non-controlled trials, nine studies showed significant improvements in weight at follow-ups between 2.5 months and three years (35–43). Seven studies showed improvements in BMI at follow-ups between 2.5 to six months (35, 36, 38, 39, 42–44). Four studies found significant improvements in waist circumference (36, 39, 42, 43). Four studies reported significant improvements in HbA1c-Levels (36, 37, 40, 44). The studies showed insignificant impacts on glucose levels, triglycerides, HDL-C, LDL-C and total cholesterol (36, 40, 42–44). Five studies reported significant blood pressure changes (38–40, 43, 44), and one study showed only significant changes in men but not in women (39). Further outcomes included improved physical activity frequency (38, 43) vegetable intake, caloric intake, and improved dietary habits such as adopting a Mediterranean diet (38, 39, 42, 43). Additionally, positive effects of telemedicine interventions on work absenteeism (35), self-efficacy, and knowledge regarding nutrition (38) were found. One study examined the effect of a telemedicine intervention and also assessed depression but did not find a significant effect for patients with prediabetes (40). Two studies observed that stronger intervention involvement was associated with increased effects (37, 41).

## DISCUSSION

This systematic review of interventions for T2D prevention delivered using telemedicine found a reduction in some but not all risk factors for developing T2D. Our meta-analysis found that lifestyle interventions, primarily diet and exercise delivered using telemedicine, reduced body weight, BMI, waist circumference, and Hba1c. No changes were seen in FPG, blood lipids or blood pressure. The subgroup analysis found larger effects for people with obesity/overweight. Subgroup meta-analyses supported the overall results. The degree of behavioural support did not seem to impact the effect. These findings highlight the potential for further application of telemedicine in primary care.

Previous meta-analyses have primarily focused on people with T2D or prevention of diabetes using the DPP approach. One meta-analysis of interventions, including 13 RCTs investigating the effect of the DPP using telemedicine, found a reduction in body weight (12). Like our review, the studies identified used many different telemedicine approaches, including web-based Apps, mobile phone Apps, text messages, digital video discs (DVDs), interactive voice response telephone calls, video conferencing, and video on-demand programming. In contrast to our study, they found that a higher degree of behavioural support led to a greater magnitude of weight loss, whereas this was not observed in our study. This discrepancy may be due to differences in the approaches used for behavioural support, which we were not able to group more specifically because of the heterogeneity in approaches and interventions. Another systematic review and meta-analysis, evaluating the effect of using mobile phone apps to promote weight loss, identified 12 RCTs and a reduction of body weight of -1.04 kg 95% CI: -1.75, -0.34) (45). In this systematic review and meta-analysis, we found a slightly higher decrease in body weight, but our population also had to be at a high risk of T2D, whereas Matteo et al did not have this inclusion criterion. This was particularly supported by our finding of a stronger effect among people with overweight/obesity as criterion for being at risk of T2D. A systematic review of people diagnosed with T2D and telemedicine interventions, including any type of intervention with focus beyond diet and exercise, showed a reduction in HbA1c (-0.415%, 95% CI: -0.482, - 0.348) (46). These results were in the same direction as we observed, but stronger, which may be due to the lower initial HbA1c in the participants included in our studies. Also, our review included other intervention types that focused primarily on lifestyle changes and not reduction of blood glucose specifically. While other systematic reviews of telemedicine interventions addressing participants with prediabetes have found a reduction in blood pressure, we did not observe a statistically significant reduction (47). This may be because few interventions included in our review did not focus on blood pressure and investigated changes in blood pressure as a secondary outcome.

Our findings in the subgroup meta-analysis highlighted that delivering diet and exercise interventions to prevent T2D using telemedicine was most effective in studies focused on people with overweight/obesity for reducing body weight, waist circumference, and HbA1c, but not for blood lipids or blood pressure. These results may reflect that most studies primarily targeted body weight, waist circumference and/or HbA1c. Regarding the duration of intervention, the effect seemed to be strongest for interventions lasting fewer than 12 months, whereas longer interventions tended to show no benefit. A reason for this could be that adherence to any lifestyle intervention is often reduced over time, with or without telemedicine. Hence, telemedicine may not necessarily improve adherence in the long-term but rather facilitate delivery of the intervention. Because the telemedicine interventions in this study varied in terms of using differnt combinations of telemedicine approaches (e.g., tele-education, tele-monitoring and tele-mentoring) and the technology used, we could not conclude that a specific approach was better than others.Given the current pace of technological development, for example, with large language models (48), telemedicine interventions are likely to evolve rapidly in the near future (49).

Our results suggest that there is a benefit of delivering prevention of T2D through telemedicine that could be implemented in a primary care setting. With the rising number of people at risk of T2D (1), and stretched health care systems, cost-effective prevention is of utmost importance. Telemedicine, in its many forms, offers such opportunities. Though the effect sizes observed in this study were modest, future technological improvements may improve these results, translating to stronger effect sizes. It has also been suggested that this may be particularly useful in rural areas with limited access to health care (50). While the direction of the effects is consistent, given the heterogeneity of the interventions, we urge readers to interpret the effects with caution.

In this systematic review and meta-analysis, we performed a comprehensive search in a wide range of databases, conducted analysis, and reported results according to current best practice guidelines. While similar systematic reviews on the topic only evaluated study quality, this study also assessed the risk of bias (12). Because there was a small number of included studies, we used a Bayesian meta-analysis approach, though results were largely similar compared with inverse variance weighted meta-analysis. Our study also included multiple risk factors instead of only investigating body weight (12).

A limitation of our study is that the search was not updated after December 2020. Many recent studies were performed during the Covid-19 pandemic or use newer technologies, which may be difficult to compare with the studies already included in the review. We did not obtain complete information on results to perform a meta-analysis for all eligible RCTs because several authors did not respond to provide the necessary data that was requested. Lastly, some publication bias was identified in sensitivity analyses. These sensitivity analyses show that the effect on body weight may be attenuated due to publication bias, but still shows a reduction.

In conclusion, our systematic evaluation highlights a clear potential for telemedicine-delivered interventions in primary care settings to prevent T2D in people with high risk of developing T2D. The effects were relatively strong among people with overweight/obesity, though the effect sizes in general were modest. The use of telemedicine may improve outcomes for people at high risk of T2D in primary care. Given the current technological advances, telemedicine today will likely not be the telemedicine of tomorrow. Future interventions may benefit from investigating how telemedicine can facilitate long-term adherence to interventions.

## Data Availability

All data produced in the present study are available upon reasonable request to the authors

## Acknowledgments

This work was initiated as part of the Global Diabetes Journal Club, an open online journal club for researchers interested in diabetes. We would like to acknowledge all those who participated in the meetings and those part of the idea generation in the working groups. A special thanks to Enzo Cerullo for commenting on the protocol and librarian Selina Lock from Leicester University for assistance with the systematic searches.

