## Supplemental for "The effectiveness of telemedicine in the prevention of type 2 diabetes: a systematic review and meta-analysis of interventions"

[1]Professorship of Public Health and Prevention, TUM School of Medicine and Health, Technical University of Munich, Germany; [2]Mathematica, Princeton, New Jersey, United States; [3]MRC Epidemiology Unit, University of Cambridge School of Clinical Medicine, Cambridge, United Kingdom; [4]Faculty of Medicine and Biomedical Sciences, University of Yaoundé 1, Yaoundé, Cameroon; [5]Department of Clinical Sciences Malmö, Lund University, Malmö, Sweden; [6]Universidad Peruana Cayetano Heredia, Lima, Perú; [7]Clínica San Felipe, Lima, Peru; [8]School of Health, University of the Sunshine Coast, Petrie, Australia; [9]Department of Medicine, University of Melbourne, Parkville, Australia; [10]School of Medicine, Western Sydney University, Campbelltown, Australia; [11]Leicester Real World Evidence Unit, Diabetes Research Centre, University of Leicester, Leicester, United Kingdom; [12]Department of Public Health, Aarhus University, Aarhus, Denmark; [13]Steno Diabetes Center Aarhus, Aarhus University Hospital, Aarhus, Denmark; [14]Department of Nutrition, Sports and Exercise, University of Copenhagen, Frederiksberg, Denmark

Supplemental Table 4. Subgroup meta-analysis of randomized controlled trials delivering lifestyle interventions using telemedicine to reduce body mass index9

Supplemental Table 8. Subgroup meta-analysis of randomized controlled trials delivering lifestyle interventions using telemedicine to reduce LDL cholesterol 17

**Supplemental Tables**

### Supplemental Table 1. Search terms for Medline[1]

**Population**

1. glycated hemoglobin A [mh]
2. hbA1C.mp.
3. blood glucose [mh]
4. blood sugar.mp.
5. glycaemia.mp.
6. glycemia.mp.
7. prediabet*.mp.
8. fasting blood glucose.mp.
9. fasting glycemia.mp.
10. fasting glycaemia.mp.
11. postprandial blood glucose.mp.
12. post-prandial blood glucose.mp.
13. postprandial glycemia.mp.
14. postprandial glycaemia.mp.
15. glucose intolerance [mh]
16. glucose intolerance.mp.
17. glucose tolerance.mp.
18. impaired glucose.mp.
19. IGT.mp.
20. impaired glycaemia.mp.
21. impaired glycemia.mp.
22. impaired fasting glucose.mp.
23. impaired fasting glyc*.mp.
24. high blood sugar.mp.
25. abnormal blood glucose.mp.
26. elevated blood glucose.mp.
27. abnormal glycemia.mp.
28. elevated glycemia.mp.
29. abnormal glycaemia.mp.
30. elevated glycaemia.mp.
31. dysglycaemia.mp.
32. dysglycemia.mp.
33. intermediate hyperglycaemia.mp.
34. intermediate hyperglycemia.mp.
35. Hyperglycaemia.mp.
36. Hyperglycemia.mp.
37. Hyperglycemia [mh]
38. insulin resistance [mh]
39. insulin resistance.mp.
40. insulin sensitivity.mp
44. non diabet*.mp.
45. non-diabet*.mp.
46. nondiabet*.mp.
47. exp Diabetes Mellitus, Type 2/
48. Diabetes mellitus [mh]
49. Diabetes mellitus.mp.
50. diabetes.mp.
51. Type 2 adj5 diabet*.mp.
52. type 2 diabet*.mp.
53. type ii diabet*.mp.
54. diabetes type 2.mp.
55. diabetes type ii.mp.
56. diabetes mellitus type 2.mp.
57. diabetes mellitus type ii.mp.
58. non-insulin dependent diabetes.mp.
59. non-insulin-dependent diabetes mellitus.mp.
60. noninsulin dependent diabetes mellitus.mp.
61. noninsulin-dependent diabetes mellitus.mp.
62. non insulin dependent diabetes mellitus.mp.
63. T2DM.mp.
64. T2D.mp.
65. metabolic syndrome [mh]
66. 1 or 2…or 65

41. hyperinsulinism [mh]
42. hyperinsulinaemia.mp.
43. Hyperinsulinemia.mp.

**Intervention**

67. Telemedicine [mh]
68. Mobile Applications [mh]
69. Cell Phone [mh]
70. Text Messaging [mh]
71. internet-Based Intervention [mh]
72. “Internet of Things” [mh]
73. internet [mh]
74. Telemedicine.mp.
75. ehealth.mp.
76. mhealth.mp.
77. e-health.mp.
78. m-health.mp.
79. mobile health.mp.
80. Telehealth.mp.
81. mobile phone.mp.
82. electronic health.mp.
83. Electronic messaging.mp.
84. email.mp.
85. Mobile app*.mp.
86. telehealth [exp]
87. internet.mp.
88. “text messag*”.mp.
89. 67 or 68… or 88

**Prevention**

90. “primary Prevention” [mh]
91. prevent*.mp.
92. control.mp.
93. Management.mp.
94. “Risk Reduction Behavior” [mh]
95. risk reduction.mp.
96. lifestyle modification*.mp.
97. “Life style”[mh]
98. life style.mp.
99. lifestyle.mp
100. lifestyle intervention.mp.
101. lifestyle advice.mp.
102. lifestyle education*.mp.
103. lifestyle discuss*.mp.
104. lifestyle change*.mp.
105. lifestyle counsel*.mp.
106. lifestyle factor*.mp.
107. behav*.mp
108. behav* intervention.mp.
109. behav* advice.mp.
110. behav* educat*.mp.
111. behav* discuss*.mp.
112. behav* modif*.mp.
113. behav* chang*.mp.
121. diet [mh]
122. “diet, healthy” [mh]
123. “Healthy lifestyle” [mh]
124. Exercise[mh]
125. Physical activity.mp.
126. exercis*.mp.
127. weight loss.mp.
128. Weight Loss [mh]
129. weight reduc*.mp.
130. weight change*.mp.
131. Body Mass Index [mh]
132. bmi.mp.
133. Metformin [mh]
134. metformin.mp.
135. Obesity[mh]
136. Obesity.mp.
137. Overweight[mh]
138. 90 or 91…or 137

114. behav* counsel*.mp.
115. Sedentary Behavior [mh]
116. habits [mh]
117. habit*.mp.
118. diet*.mp.
119. nutrition*.mp.
120. weight management.mp.

**Primary Health Care**

139. Primary Health Care [mh]
140. primary care [mh]
141. primary health care [mh]
142. primary medical care.mp.
143. medical care.mp.
144. family practice[mh]
145. Family practic*.mp.
146. family doctor*.mp.
147. family medicine.mp.
148. general practice*.mp.
149. physician*.mp.
150. pharmac*.mp.
151. doctor*.mp.
152. nurs*.mp.
153. health professional*
154. GP.mp.
155. general practitioner*.mp.
156. practitioner*.mp.
157. Physicians, Primary Care [mh]
158. Primary Care Nursing[mh]
159. Nutritionists [mh]
160. nutritionist*.mp.
161. Physical Therapists [mh]
162. Nurses [mh]
163. dietitian [mh]
164. physiotherapist [mh]
165. general practitioners[mh]
166. General practitioner*.mp.
167. prevent* medicine.mp.
168. 139 or 140…or 167

**Combination of all terms**

169. 66 and 89 and 138 and 168

---

Supplemental Table 2. Overview of PICO-S criteria for systematic review

| | |
|---|---|
| Population | - Adults aged ≥18 years<br>- Adults at high risk of developing type 2 diabetes, e.g. prediabetes diagnosis, metabolic syndrome, overweight/obesity, and/or prediabetes screening tools |

| | |
|---|---|
| Intervention | *Primary prevention:*<br><br>- Lifestyle Interventions targeting physical activity and/or diet<br>- Pharmacological (metformin)<br><br>*Technologies*<br><br>- Tele-education, telemonitoring, telecase-management, telementoring and teleconsultation<br>- Other technologically assisted (including but not limited to the use of mobile phone or tablet) |
| Comparator | - No restrictions |
| Outcome | For inclusion in meta-analysis:<br>- metabolic syndrome and/or its individual components including:<br>i) changes in adiposity (body weight, body mass index, waist circumference)<br>ii) incidence of type 2 diabetes<br>iii) changes in markers of glucose homeostasis (fasting blood glucose, HbA1c, 2-hour glycaemia, HOMA-IR)<br>iv) changes in blood lipids (LDL, HDL, TG)<br>v) changes in blood pressure (SBP, DBP)<br><br>For inclusion in narrative synthesis:<br>- No restriction |
| Study design | - Quantitative interventional studies containing empirical data (primary or secondary)<br>- Studies in all countries<br>- The intervention is relevant to or takes place in a primary health care setting:<br> - Primary health care refers to the first point of contact with the health system and can be provided in the home or in community-based settings such as general practices, other private medical practices, community health centres, local government, and non-government service settings<br> - Through general practitioner (GP), nurses, allied health professionals, midwives, pharmacists, dentists etc. |

Supplemental Table 3. Overview of risk of bias (RoB) from RCTs, including individual components

| Author, year | Sequence generation | Allocation concealment | Blinding of participants and personnel | Blinding of outcome assessment | Incomplete outcome data | Selective reporting | Other sources of bias | Overall risk of bias |
|---|---|---|---|---|---|---|---|---|
| Aguiar, 2016 | Low | Low | Unsure | Low | Low | Low | Unsure | Low |
| Allman-Farinelli, 2016 | Low | Unsure | Unsure | Low | Unsure | Unsure | High | High |
| Apianiz, 2019 | Low | Unsure | Low | Unsure | Unsure | Low | Unsure | Some |
| Bender, 2018 | Low | Low | Unsure | Low | Low | Low | Unsure | Some |
| Block, 2015 | Low | Low | Low | Unsure | Low | Low | Unsure | Low |
| Bosak, 2010 | Unsure | Unsure | Unsure | Unsure | Low | Low | Unsure | High |
| Carnie, 2013 | Low | Low | Unsure | Low | Low | Low | Unsure | Low |
| Cho, 2020 | Low | Low | Low | Low | Unsure | Low | Unsure | Low |
| Cicolini, 2014 | Low | Low | Low | Unsure | Low | Low | Low | Low |
| Chen, 2020 | Low | Low | Low | Low | Some | Some | Low | Some |
| Fischer, 2016 | Low | Unsure | High | High | High | Low | Low | High |
| Fukuoka, 2015 | Low | High | Unsure | Unsure | Low | Low | Low | High |
| Green, 2014 | Low | Low | Unsure | Unsure | Low | Unsure | Low | Some |
| Hebden, 2013 | Low | Low | Unsure | Unsure | Low | Unsure | Low | Some |
| Hurkmans, 2018 | Low | Low | Low | Low | High | High | Low | High |
| Jahangiry, 2015 | Low | Unsure | Unsure | Unsure | Low | Low | Unsure | High |
| Johnson, 2019 | Low | Low | Low | Low | Low | Low | Unsure | Low |
| Kempf, 2018 | Low | Low | Low | Low | Low | Low | Low | Low |

| | | | | | | | | |
|---|---|---|---|---|---|---|---|---|
| Lison, 2020 | Low | Low | Low | Low | Low | Low | Unsure | Low |
| Ma, 2013 | Low | Low | Low | Low | Low | Low | Low | Low |
| McLeod, 2020 | Low | Low | Low | Low | Low | Low | Low | Low |
| Parez Ewert, 2016 | Low | Low | Unsure | Unsure | Low | Low | Low | Low |
| Patel, 2019 | Low | Low | Low | Unsure | High | Low | Low | High |
| Peyer, 2017 | Low | Low | Some | Unsure | Unsure | Unsure | Unsure | Some |
| Rogers, 2016 | Unsure | Low | Unsure | Unsure | High | Low | Low | High |
| Silina, 2017 | Low | Unsure | Unsure | Unsure | Low | Unsure | Low | Some |
| Staite, 2020 | Low | Low | Low | Low | High | Low | Low | High |
| Tanaka, 2018 | Low | Low | Low | Low | Unsure | Low | Low | Low |
| Tarraga Marcos, 2017 | Low | Low | Unsure | Unsure | Low | Unsure | Low | Some |
| Toro-Ramos, 2020 | Low | Low | Low | Unsure | Unsure | Unsure | High | High |
| Weinstock, 2013 | Low | Unsure | Low | Unsure | High | Low | Unsure | High |
| Wu, 2011 | Low | High | High | Unsure | High | Unsure | Low | High |

.

Supplemental Table 4. Subgroup meta-analysis of randomized controlled trials delivering lifestyle interventions using telemedicine to reduce body mass index

| Subgroup | N RCTs | MD (95% CI) | $I^2$ |
|---|---|---|---|
| **Overall** | 11 | -0.71 (-1.06, -0.37) | 70% |
| **Type of control group** | | | |
| Intervention without telemedicine | 2 | -0.77 (-1.32, -0.22) | 25% |
| Usual care | 3 | -0.44 (-0.85, -0.03) | 12% |
| Minimal telemedicine | 2 | -0.37 (-0.98, 0.24) | 0% |
| Wait list control | 4 | -1.04 (-1.41, -0.66) | 65% |
| **Degree of behavioral support** | | | |
| No support | 3 | -0.98 (-1.57, -0.39) | 88% |
| Supported by remote contact | 6 | -0.62 (-1.02, -0.21) | 0% |
| Supported by face-to-face contact | 1 | -0.8 (-2.78, 0.18) | - |
| Supported by remote and face-to-face contact | 1 | -0.50 (-1.38, 0.38) | - |
| **Type of lifestyle intervention** | | | |
| Diet | 2 | -0.37 (-0.96, 0.21) | 0% |
| Diet and exercise | 9 | -0.79 (-1.08, 0.50) | 58% |
| **Population at high-risk** | | | |
| Overweight/obese | 6 | -0.75 (-1.19, -0.32) | 76% |
| Prediabetes | 2 | -0.54 (-1.21, 0.13) | 0% |
| Prediabetes and overweight/obese | 2 | -0.82 (-1.54, -0.09) | 0% |
| Metabolic syndrome | 1 | -0.70 (-1.70, 0.30) | - |
| **Risk of bias** | | | |
| Low | 2 | -1.24 (-1.81, -0.68) | 85% |
| Moderate | 4 | -0.61 (-0.98, -0.23) | 54% |

| | | | |
|---|---|---|---|
| High | 5 | -0.58 (-0.93, -0.23) | 0% |
| **Duration of intervention** | | | |
| Short (<6 months) | 5 | -0.57 (-0.97, -0.18) | 23% |
| Medium (6 to <12 months) | 3 | -0.92 (-1.46, -0.38) | 85% |
| Long (>= 12 months) | 3 | -0.76 (-1,29, -0.23) | 0% |

Supplemental Table 5. Subgroup meta-analysis of randomized controlled trials delivering lifestyle interventions using telemedicine to reduce waist circumference

| Subgroup | N RCTs | MD (95% CI) | $I^2$ |
|---|---|---|---|
| **Overall** | 8 | -2.70 (-4.62, -0.74) | 70% |
| **Type of control group** | | | |
| Intervention without telemedicine | 1 | -4.60 (-6.85, -2.35) | - |
| Usual care | 3 | -2.09 (-.02, -1.17) | 0% |
| Usual care and minimal telemedicine | 1 | -0.60 (-2.46, 1.26) | - |
| Wait list control | 3 | -4.43 (-5.88, -3.00) | 73% |
| **Degree of behavioral support** | | | |
| No support | 3 | -3.46 (-5.74, -1.17) | 85% |
| Supported by remote contact | 5 | -2.24 (-4.07, -0.41) | 49% |
| Supported by remote and face-to-face contact | - | - | - |
| **Type of lifestyle intervention** | | | |
| Overall behaviour | - | - | - |
| Diet | 1 | -1.70 (-5.45, 2.05) | - |
| Diet and exercise | 7 | -2.89 (-4.47, -1.32) | 71% |
| **Population at high-risk** | | | |
| Overweight/obese | 2 | -5.00 (-7.03, -2.97) | 0% |
| Prediabetes | - | - | - |
| Prediabetes and overweight/obese | 3 | -2.51 (-4.07, -0.95) | 74% |
| Metabolic syndrome | 3 | -1.35 (-3.08, 0.39) | 0% |
| **Risk of bias** | | | |
| Low | 4 | -2.79 (-4.13, -1.46) | 67% |
| Moderate | 2 | -4.74 (-6.93, -2.56) | 0% |

| | | | |
|---|---|---|---|
| High | 2 | -0.19 (-2.54, 2.17) | 0% |
| **Duration of intervention** | | | |
| Short (<6 months) | 2 | -3.15 (-6.40, 0.09) | 80% |
| Medium (6 to <12 months) | 3 | -2.51 (-5.39, 0.36) | 80% |
| Long (>= 12 months) | 3 | -2.56 (-5.19, 0.07) | 73% |

Supplemental Table 6. Subgroup meta-analysis of randomized controlled trials delivering lifestyle interventions using telemedicine to reduce fasting plasma glucose

| **Subgroup** | **N RCTs** | **MD (95% CI)** | **$I^2$** |
|---|---|---|---|
| **Overall** | 9 | -0.05 (-0.19, 0.11) | 0% |
| **Type of control group** | | | |
| Intervention without telemedicine | 1 | -0.01 (-0.25, 0.23) | - |
| Usual care | 5 | -0.05 (-0.19, 0.09) | 11% |
| Usual care and minimal telemedicine | - | - | - |
| Wait list control | 3 | -0.07 (-0.27, 0.12) | 0% |
| **Degree of behavioral support** | | | |
| No support | 2 | -0.02 (-0.17, 0.12) | 0% |
| Supported by remote contact | 5 | -0.11 (-0.20, -0.01) | 0% |
| Supported by remote and face-to-face contact | 1 | -0-02 (-1.22, 1.18) | - |
| Supported by face-to-face contact | 1 | 0.13 (-0.13, 0.39)) | - |
| **Type of lifestyle intervention** | | | |
| Overall behaviour | - | - | - |
| Diet | 2 | 0.02 (-0.21, 0.25) | 0% |
| Diet and exercise | 7 | -0.07 (-0.17, 0.03) | 0% |
| **Population at high-risk** | | | |
| Overweight/obese | 3 | -0.01 (-0.14, 0.12) | 0% |
| Prediabetes | 1 | -0.02 (-1.22, 1.18) | - |
| Prediabetes and overweight/obese | 3 | -0.10 (-0.21, 0.02)) | 48% |
| Metabolic syndrome | 2 | -0.10 (-0.51, 0.31) | 0% |
| **Risk of bias** | | | |
| Low | 3 | -0.14 (-0.25, -0.03) | 0% |

| | | | |
|---|---|---|---|
| Moderate | 4 | -0.01 (-0.14, 0.11) | 0% |
| High | 2 | 0.06 (-0.16, 0.28) | 0% |
| **<u>Duration of intervention</u>** | | | |
| Short (<6 months) | 3 | 0.02 (-0.17, 0.21) | 0% |
| Medium (6 to <12 months) | 4 | -0.02 (-0.18, 0.14) | 0% |
| Long (>= 12 months) | 2 | -0.11 (-0.22, 0.01) | 46% |

Supplemental Table 7. Subgroup meta-analysis of randomized controlled trials delivering lifestyle interventions using telemedicine to reduce HbA1c

| Subgroup | N RCTs | MD (95% CI) | $I^2$ |
|---|---|---|---|
| **Overall** | 11 | -0.07 (-0.14, 0.00) | 71% |
| **Type of control group** | | | |
| Intervention without telemedicine | 2 | -0.06 (-0.15, 0.03) | 29% |
| Usual care | 6 | -0.07 (-0.13, -0.01) | 31% |
| Usual care and minimal telemedicine | 1 | 0.04 (-0.05, 1.14) | - |
| Wait list control | 2 | -0.15 (-0.24, -0.06) | 45% |
| **Degree of behavioral support** | | | |
| No support | 2 | -0.06 (-0.19, 0.05) | 93% |
| Supported by remote contact | 7 | -0.08 (-0.16, -0.01) | 11% |
| Supported by remote and face-to-face contact | 1 | -0.10 (-0.30, 0.10) | - |
| Supported by face-to-face contact | 1 | 0.07 (-0.18, 0.32) | - |
| **Type of lifestyle intervention** | | | |
| Overall behaviour | 1 | 0.00 (-0.17, 0.17) | - |
| Diet | 3 | -0.11 (-0.21, -0.00) | 0% |
| Diet and exercise | 7 | -0.07 (-0.14, 0.00) | 74% |
| **Population at high-risk** | | | |
| Overweight/obese | 3 | -0.13 (-0.21, -0.05) | 28% |
| Prediabetes | 3 | -0.03 (-0.12, 0.06) | 0% |
| Prediabetes and overweight/obese | 4 | -0.02 (-0.11, 0.06) | 69% |
| Metabolic syndrome | 1 | -0.12 (-0.28, 0.03) | - |
| **Risk of bias** | | | |
| Low | 3 | -0.11 (-0.18, -0.04) | 74% |

| | | | |
|---|---|---|---|
| Moderate | 5 | -0.11 (-0.17, -0.05) | 0% |
| High | 3 | 0.04 (-0.03, 0.11) | 0% |
| **<u>Duration of intervention</u>** | | | |
| Short (<6 months) | 4 | -0.09 (-0.15, -0.03) | 0% |
| Medium (6 to <12 months) | 3 | -0.13 (-0.19, -0.07) | 28% |
| Long (>= 12 months) | 4 | 0.02 (-0.03, 0.07) | 41% |

Supplemental Table 8. Subgroup meta-analysis of randomized controlled trials delivering lifestyle interventions using telemedicine to reduce LDL cholesterol

| **Subgroup** | **N RCTs** | **MD (95% CI)** | **$I^2$** |
|---|---|---|---|
| **Overall** | 8 | -0.04 (-0.17, 0.09) | 40% |
| **Type of control group** | | | |
| Intervention without telemedicine | 1 | -0.02 (-0.22, 0.18) | - |
| Usual care | 5 | -0.05 (-0.16, 0.05) | 0% |
| Usual care and minimal telemedicine | 1 | 0.02 (-0.19, 0.23) | - |
| Wait list control | 1 | -0.10 (-0.34, 0.14) | - |
| **Degree of behavioral support** | | | |
| No support | 2 | -0.00 (-0.14, 0.14 ) | 0% |
| Supported by remote contact | 4 | -0.09 (-0.19, 0.02) | 0% |
| Supported by remote and face-to-face contact | 1 | -0.25 (-0.80, 0.29) | - |
| Supported by face-to-face contact | 1 | 0.15 (-0.11, 0.41) | - |
| **Type of lifestyle intervention** | | | |
| Overall behaviour | - | - | - |
| Diet | 3 | 0.01 (-0.12, 0.15) | 0% |
| Diet and exercise | 5 | -0.07 (-0.17, 0.03) | 0% |
| **Population at high-risk** | | | |
| Overweight/obese | 2 | -0.03 (-0.19, 0.12) | 0% |
| Prediabetes | 1 | 0.15 (-0.11, 0.41) | - |
| Prediabetes and overweight/obese | 3 | -0.08 (-0.21, 0.04) | 0% |
| Metabolic syndrome | 2 | -0.06 (-0.22, 0.09) | 0% |
| **Risk of bias** | | | |
| Low | 2 | -0.10 (-0.23, 0.04) | 0% |

| | | | |
|---|---|---|---|
| Moderate | 3 | 0.02 (-0.12, 0.15) | 0% |
| High | 3 | -0.05 (-0.20, 0.10) | 0% |
| **Duration of intervention** | | | |
| Short (<6 months) | 3 | 0.01 (-0.14, 0.17) | 9% |
| Medium (6 to <12 months) | 2 | -0.07 (-0.25, 0.10) | 0% |
| Long (>= 12 months) | 3 | -0.06 (-0.17, 0.05) | 0% |

Supplemental Table 9. Subgroup meta-analysis of randomized controlled trials delivering lifestyle interventions using telemedicine to reduce HDL cholesterol

| **Subgroup** | **N RCTs** | **MD (95% CI)** | **$I^2$** |
|---|---|---|---|
| **Overall** | 9 | 0.02 (-0.02, 0.07) | 18% |
| **Type of control group** | | | |
| Intervention without telemedicine | 1 | 0.14 (-0.23, 0.51) | - |
| Usual care | 6 | -0.00 (-0.00, 0.00) | 0% |
| Usual care and minimal telemedicine | 1 | 0.00 (-0.07, 0.07) | - |
| Wait list control | 1 | 0.06 (0.01, 0.11) | - |
| **Degree of behavioral support** | | | |
| No support | 2 | 0.01 (-0.07, 0.08) | 0% |
| Supported by remote contact | 5 | 0.02 (-0.01, 0.05) | 45% |
| Supported by remote and face-to-face contact | 1 | 0.06 (-0.04, 0.16) | - |
| Supported by face-to-face contact | 1 | -0.04 (-1.20, 1.10) | - |
| **Type of lifestyle intervention** | | | |
| Overall behaviour | - | - | - |
| Diet | 3 | 0.05 (-0.01, 0.10) | 0% |
| Diet and exercise | 6 | 0.01 (-0.02, 0.03) | 21% |
| **Population at high-risk** | | | |
| Overweight/obese | 2 | 0.05 (-0.04, 0.15) | 0% |
| Prediabetes | 1 | 0.06 (-0.04, 0.16) | - |
| Prediabetes and overweight/obese | 3 | -0.00 (-0.06, 0.05) | 0% |
| Metabolic syndrome | 3 | 0.02 (-0.02, 0.06) | 65% |
| **Risk of bias** | | | |
| Low | 3 | -0.00 (-0.00, 0.00) | 0% |

| | | | |
|---|---|---|---|
| Moderate | 3 | 0.06 (-0.00, 0.11) | 0% |
| High | 3 | 0.04 (-0.00, 0.08) | 4% |
| **Duration of intervention** | | | |
| Short (<6 months) | 3 | 0.04 (-0.03, 0.12) | 0% |
| Medium (6 to <12 months) | 3 | 0.02 (-0.01, 0.06) | 72% |
| Long (>= 12 months) | 3 | -0.00 (-0.07, 0.05) | 0% |

Supplemental Table 10. Subgroup meta-analysis of randomized controlled trials delivering lifestyle interventions using telemedicine to reduce triglycerides

| Subgroup | N RCTs | MD (95% CI) | $I^2$ |
|---|---|---|---|
| **Overall** | 8 | 0.00 (-0.24, 0.26) | 92% |
| **Type of control group** | | | |
| Intervention without telemedicine | 1 | 0.23 (-0.11, 0.57) | - |
| Usual care | 5 | -0.08 (-0.22, 0.05) | 37% |
| Usual care and minimal telemedicine | 1 | 0.39 (-0.06, 0.84) | - |
| Wait list control | 1 | 0.02 (-0.18, 0.22) | - |
| **Degree of behavioral support** | | | |
| No support | 2 | 0.28 (0.04, 0.52) | 0% |
| Supported by remote contact | 4 | -0.01 (-0.01, 0.00) | 0% |
| Supported by remote and face-to-face contact | 1 | -0.36 (-0.68, -0.04) | - |
| Supported by face-to-face contact | 1 | -0.04 (-0.46, 0.37) | - |
| **Type of lifestyle intervention** | | | |
| Overall behaviour | - | - | - |
| Diet | 2 | -0.11 (-0.36, 0.12) | 79% |
| Diet and exercise | 6 | 0.06 (-0.12, 0.23) | 35% |
| **Population at high-risk** | | | |
| Overweight/obese | 1 | 0.23 (-0.06, 0.52) | - |
| Prediabetes | 1 | -0.36 (-0.68, -0.04) | - |
| Prediabetes and overweight/obese | 3 | -0.04 (-0.19, 0.10) | 59% |
| Metabolic syndrome | 3 | -0.01 (-0.01, 0.00) | 0% |
| **Risk of bias** | | | |
| Low | 3 | -0.05 (-0.32, 0.21) | 0% |

| | | | |
|---|---|---|---|
| Moderate | 2 | -0.05 (-0.39, 0.28) | 86% |
| High | 3 | 0.09 (-0.18, 0.36) | 35% |
| **Duration of intervention** | | | |
| Short (<6 months) | 3 | -0.11 (-0.38, 0.15) | 58% |
| Medium (6 to <12 months) | 2 | 0.02 (-0.36, 0.39) | 0% |
| Long (>= 12 months) | 3 | 0.12 (-0.15, 0.38) | 74% |

Supplemental Table 11. Subgroup meta-analysis of randomized controlled trials delivering lifestyle interventions using telemedicine to reduce systolic blood pressure

| **Subgroup** | **N RCTs** | **MD (95% CI)** | **$I^2$** |
|---|---|---|---|
| **Overall** | 10 | -2.06 (-4.38, 0.15) | 58% |
| **Type of control group** | | | |
| Intervention without telemedicine | 1 | 2.30 (-4.39, 9.00) | - |
| Usual care | 6 | -2.23 (-4.80, 0.34) | 60% |
| Usual care and minimal telemedicine | 1 | -2.62 (-8.30, 3.06) | - |
| Wait list control | 2 | -3.00 (-6.87, 0.86) | 0% |
| **Degree of behavioral support** | | | |
| No support | 2 | -2.79 (-6.48, 0.90) | 0% |
| Supported by remote contact | 7 | -1.51 (-3.42, 0.40) | 54% |
| Supported by remote and face-to-face contact | 1 | -8.40 (-16.64, -0.16) | - |
| **Type of lifestyle intervention** | | | |
| Overall behaviour | - | - | - |
| Diet | 2 | -2.90 (-7.31, 1.51) | 0% |
| Diet and exercise | 8 | -1.91 (-3.81, -0.01) | 57% |
| **Population at high-risk** | | | |
| Overweight/obese | 3 | -1.11 (-5.17, 2.95) | 23% |
| Prediabetes | - | - | - |
| Prediabetes and overweight/obese | 4 | -3.31 (-6.48, -0.17) | 63% |
| Metabolic syndrome | 3 | -1.31 (-4.89, 2.26) | 64% |
| **Risk of bias** | | | |
| Low | 4 | -0.84 (-3.41, 1.73) | 39% |
| Moderate | 3 | -2.32 (-5.91, 1.27) | 65% |

| | | | |
|---|---|---|---|
| High | 3 | -3.58 (-6.62, -0.54) | 0% |
| **Duration of intervention** | | | |
| Short (<6 months) | 2 | -4.86 (-9.88, 0.15) | 29% |
| Medium (6 to <12 months) | 5 | -0.90 (-3.67, 1.87) | 50% |
| Long (>= 12 months) | 3 | -2.53 (-5.64, 0.58) | 62% |

Supplemental Table 12. Subgroup meta-analysis of randomized controlled trials delivering lifestyle interventions using telemedicine to reduce diastolic blood pressure

| **Subgroup** | **N RCTs** | **MD (95% CI)** | **$I^2$** |
|---|---|---|---|
| **<u>Overall</u>** | 8 | -1.82 (-4.05, 0.18) | 64% |
| **<u>Type of control group</u>** | | | |
| Intervention without telemedicine | - | - | - |
| Usual care | 5 | -1.83 (-4.20, 0.54) | 52% |
| Usual care and minimal telemedicine | 1 | -1.61 (-6.37, 3.15) | - |
| Wait list control | 2 | -2.33 (-6.08, 1.41) | 77% |
| **<u>Degree of behavioral support</u>** | | | |
| No support | 2 | -0.94 (-3.21, 1.33) | 0% |
| Supported by remote contact | 5 | -1.51 (-3.05, 0.04) | 42% |
| Supported by remote and face-to-face contact | 1 | -6.5 (-11.17, -1.82) | - |
| **<u>Type of lifestyle intervention</u>** | | | |
| Overall behaviour | - | - | - |
| Diet | 2 | -2.16 (-5.60, 1.27) | 0% |
| Diet and exercise | 6 | -1.75 (-3.50, -0.00) | 61% |
| **<u>Population at high-risk</u>** | | | |
| Overweight/obese | 2 | -0.87 (-4.58, 2.82) | 0% |
| Prediabetes | - | - | - |
| Prediabetes and overweight/obese | 3 | -2.34 (-5.08, 0.38) | 67% |
| Metabolic syndrome | 3 | -2.05 (-5.08, 0.98) | 66% |
| **<u>Risk of bias</u>** | | | |
| Low | 4 | -0.58 (-2.15, 1.00) | 0% |
| Moderate | 1 | -2.00 (-6.38, 2.38 ) | - |

| | | | |
|---|---|---|---|
| High | 3 | -3.55 (-5.65, -1.45) | 60% |
| **Duration of intervention** | | | |
| Short (<6 months) | 4 | -1.36 (-3.60, 0.88) | 56% |
| Medium (6 to <12 months) | 2 | -4.10 (-7.46, -0.74) | 55% |
| Long (>= 12 months) | 2 | -1.15 (-3.54, 1.24) | 0% |

Supplemental Table 13. Bayesian meta-analysis estimates using alternative prior distributions

| Risk factor | Half-Cauchy (1.0) | Half-Cauchy (0.5) | Half-normal (1.0) | Half-normal (0.5) |
|---|---|---|---|---|
| **Measures of anthropometry** | | | | |
| Body weight, kg | -1.66 | -1.66 | -1.63 | -1.57 |
| Body mass index, kg/m$^2$ | -0.71 | -0.71 | -0.71 | -0.71 |
| Waist circumference, cm | -2.70 | -2.70 | -2.72 | -2.70 |
| **Measures of blood glucose** | | | | |
| FPG, mmol/L | -0.04 | -0.04 | -0.05 | -0.05 |
| HbA1c, % | -0.07 | -0.07 | -0.07 | -0.07 |
| **Measures of blood lipids** | | | | |
| LDL cholesterol, mmol/L | -0.04 | -0.04 | -0.04 | -0.04 |
| HDL cholesterol, mmol/L | 0.02 | 0.03 | 0.03 | 0.03 |
| TG, mmol/L | 0.00 | 0.00 | -0.00 | -0.00 |
| **Measures of blood pressure** | | | | |
| Systolic blood pressure, mmHg | -2.06 | -2.07 | -1.94 | -1.88 |
| Diastolic blood pressure, mmHg | -1.82 | -1.85 | -1.65 | -1.55 |

Supplemental Table 14. Egger's test and trim-and-fill estimates

| Risk factor | Egger's test p-value | N studies | Bayesian MD (95% CI) | N studies imputed | Trim-and-fill MD (95% CI) |
|---|---|---|---|---|---|
| **Measures of anthropometry** | | | | | |
| Body weight, kg | 0.043 | 17 | -1.66 (-2.48, -0.90) | 5 | -0.95 (-1.80, -0.11) |
| Body mass index, kg/m$^2$ | 0.456 | 11 | -0.71 (-1.06, -0.37) | 0 | -0.71 (-0.97, -0.45) |
| Waist circumference, cm | 0.824 | 8 | -2.70 (-4.62, -0.74) | 0 | -2.85 (-4.40, -1.29) |
| **Measures of blood glucose** | | | | | |
| Fasting blood glucose, mmol/L | 0.404 | 9 | -0.05 (-0.19, 0.11) | 4 | -0.12 (-0.22, -0.03) |
| HbA1c, % | 0.257 | 11 | -0.07 (-0.14, 0.00) | 1 | -0.06 (-0.12, -0.01) |
| **Measures of blood lipids** | | | | | |
| LDL cholesterol, mmol/L | 0.969 | 8 | -0.04 (-0.17, 0.09) | 0 | -0.04 (-0.12, 0.04) |
| HDL cholesterol, mmol/L | 0.860 | 9 | 0.02 (-0.02, 0.07) | 0 | 0.03 (-0.00, 0.06) |
| TG, mmol/L | 0.902 | 8 | 0.00 (-0.24, 0.26) | 0 | -0.01 (-0.01, 0.00) |
| **Measures of blood pressure** | | | | | |
| Systolic blood pressure, mmHg | 0.197 | 10 | -2.06 (-4.38, 0.15) | 3 | -1.60 (-3.15, -0.05) |
| Diastolic blood pressure, mmHg | 0.088 | 8 | -1.82 (-4.05, 0.18) | 3 | -1.01 (-2.81, 0.79) |

### Supplemental Figures

Supplemental Figure 1. Forest plots of Bayesian meta-analysis.

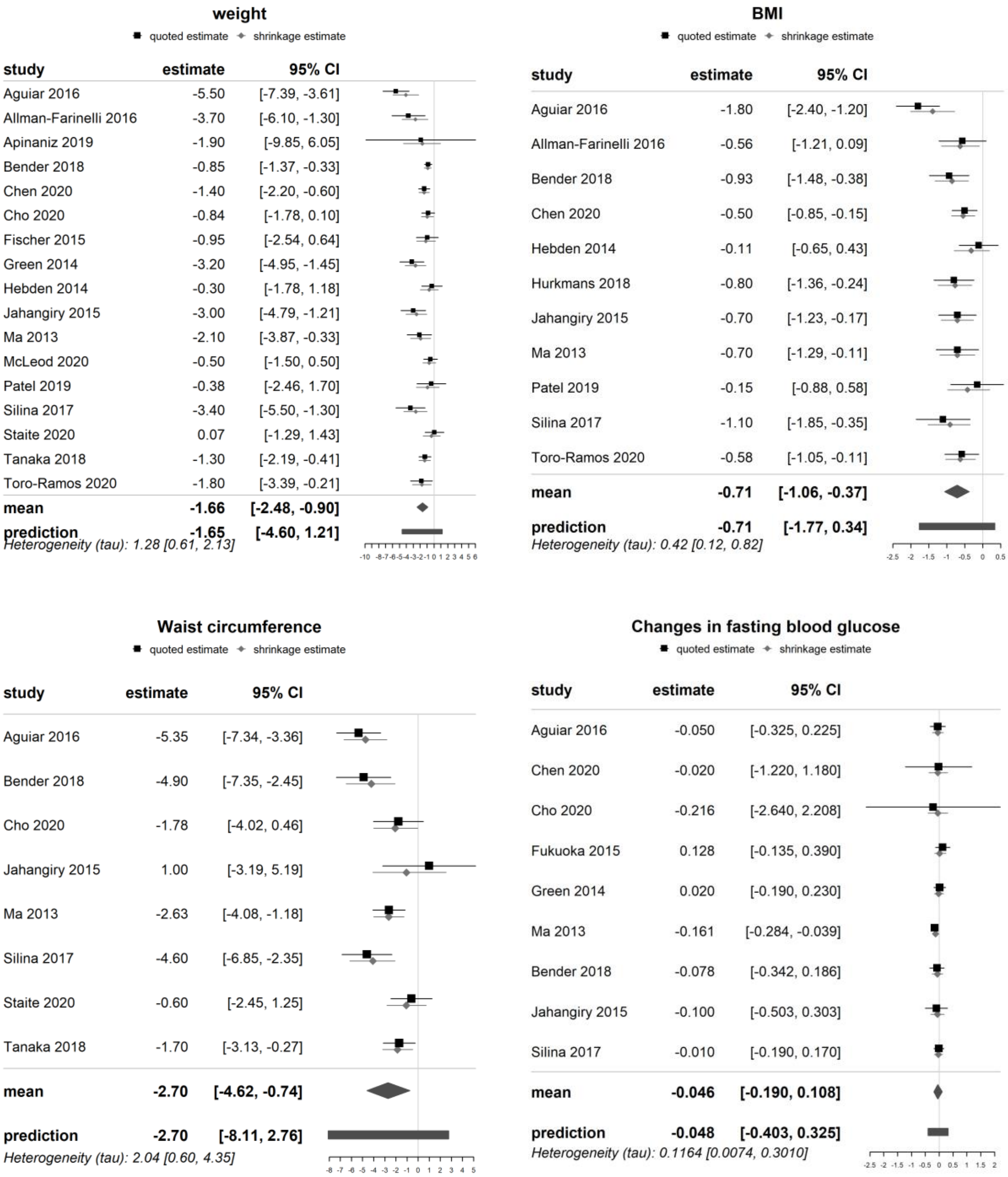

### Changes in HbA1c

■ quoted estimate ◆ shrinkage estimate

| study | estimate | 95% CI |
|---|---|---|
| Apinaniz 2019 | -0.095 | [-0.189, -0.001] |
| Toro-Ramos 2020 | 0.006 | [-0.131, 0.143] |
| Aguiar 2016 | -0.200 | [-0.300, -0.100] |
| Chen 2020 | -0.100 | [-0.225, 0.025] |
| Fischer 2015 | -0.290 | [-0.585, 0.005] |
| Fukuoka 2015 | 0.070 | [-0.129, 0.269] |
| Green 2014 | -0.100 | [-0.200, -0.000] |
| McLeod 2020 | 0.000 | [-0.100, 0.100] |
| Tanaka 2018 | -0.120 | [-0.235, -0.005] |
| Staite 2020 | 0.048 | [-0.018, 0.113] |
| Bender 2018 | -0.100 | [-0.206, 0.006] |
| **mean** | **-0.071** | **[-0.144, 0.000]** |
| **prediction** | **-0.070** | **[-0.292, 0.147]** |

*Heterogeneity (tau): 0.087 [0.031, 0.172]*

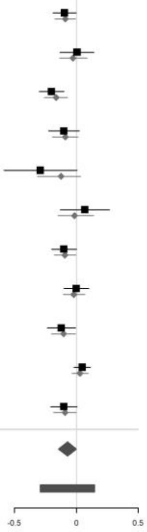

### LDL cholesterol

■ quoted estimate ◆ shrinkage estimate

| study | estimate | 95% CI |
|---|---|---|
| Chen 2020 | 0.150 | [-0.110, 0.410] |
| Fukuoka 2015 | -0.253 | [-0.796, 0.289] |
| Green 2014 | -0.047 | [-0.292, 0.199] |
| Ma 2013 | -0.140 | [-0.311, 0.032] |
| Tanaka 2018 | -0.034 | [-0.240, 0.173] |
| Staite 2020 | 0.020 | [-0.185, 0.225] |
| Jahangiry 2015 | -0.100 | [-0.339, 0.139] |
| Silina 2017 | -0.020 | [-0.220, 0.180] |
| **mean** | **-0.040** | **[-0.168, 0.087]** |
| **prediction** | **-0.041** | **[-0.350, 0.270]** |

*Heterogeneity (tau): 0.0958 [0.0040, 0.2579]*

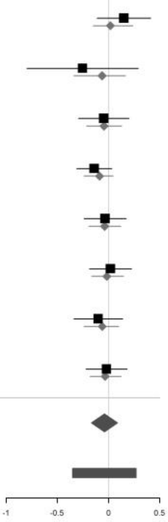

### HDL cholesterol

■ quoted estimate ◆ shrinkage estimate

| study | estimate | 95% CI |
|---|---|---|
| Chen 2020 | 0.0600 | [-0.0250, 0.1450] |
| Cho 2020 | -0.0008 | [-0.0028, 0.0013] |
| Fukuoka 2015 | -0.0440 | [-1.1887, 1.1008] |
| Green 2014 | 0.0491 | [-0.0310, 0.1293] |
| Jahangiry 2015 | 0.0600 | [0.0103, 0.1097] |
| Ma 2013 | -0.0078 | [-0.0649, 0.0493] |
| Silina 2017 | 0.1400 | [-0.2300, 0.5100] |
| Staite 2020 | 0.0000 | [-0.0650, 0.0650] |
| Tanaka 2018 | 0.0052 | [-0.1295, 0.1398] |
| **mean** | **0.0202** | **[-0.0229, 0.0705]** |
| **prediction** | **0.0191** | **[-0.0935, 0.1424]** |

*Heterogeneity (tau): 0.0383 [0.0047, 0.0990]*

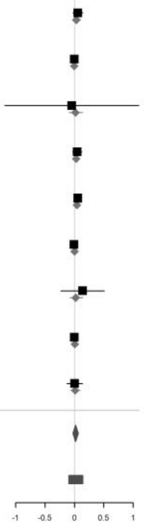

### Triglycerides

■ quoted estimate ◆ shrinkage estimate

| study | estimate | 95% CI |
|---|---|---|
| Chen 2020 | -0.3600 | [-0.6800, -0.0400] |
| Cho 2020 | 0.4209 | [-8.9699, 9.8117] |
| Fukuoka 2015 | -0.0429 | [-0.4594, 0.3736] |
| Jahangiry 2015 | 0.0200 | [-0.0560, 0.0960] |
| Ma 2013 | -0.1129 | [-0.2779, 0.0521] |
| Silina 2017 | 0.2300 | [-0.0600, 0.5200] |
| Staite 2020 | 0.3900 | [-0.0250, 0.8050] |
| Tanaka 2018 | -0.0059 | [-0.0120, 0.0002] |
| **mean** | **0.0004** | **[-0.2435, 0.2608]** |
| **prediction** | **-0.0005** | **[-0.6645, 0.6831]** |

*Heterogeneity (tau): 0.2155 [0.0066, 0.5654]*

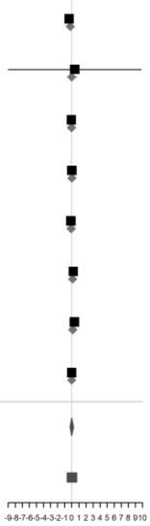

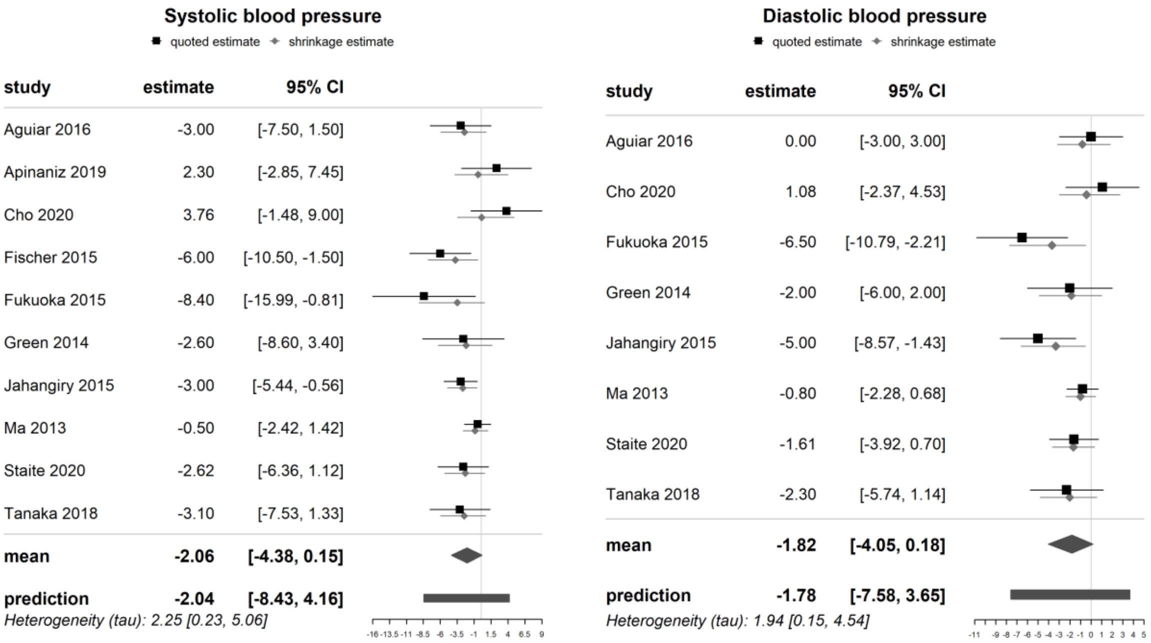

**Supplemental Figure 1.** Forest plots of Bayesian meta-analysis. Prior distribution for Tau was set using a half-Cauchy distribution and prior distribution for the mean was set to 0 with a standard deviation as the maximum value among the studies. Maximum standard deviation for body weight = 42.5, Body mass index = 5.3, waist circumference = 27.0, fasting plasma glucose = 14.0, HbA1c = 1.9, LDL = 2.2, HDL = 4.6, TG = 3.0, systolic blood pressure =30.8, diastolic blood pressure =23.0.

Supplemental Figure 2. Funnel plots with trim-and-fill studies included

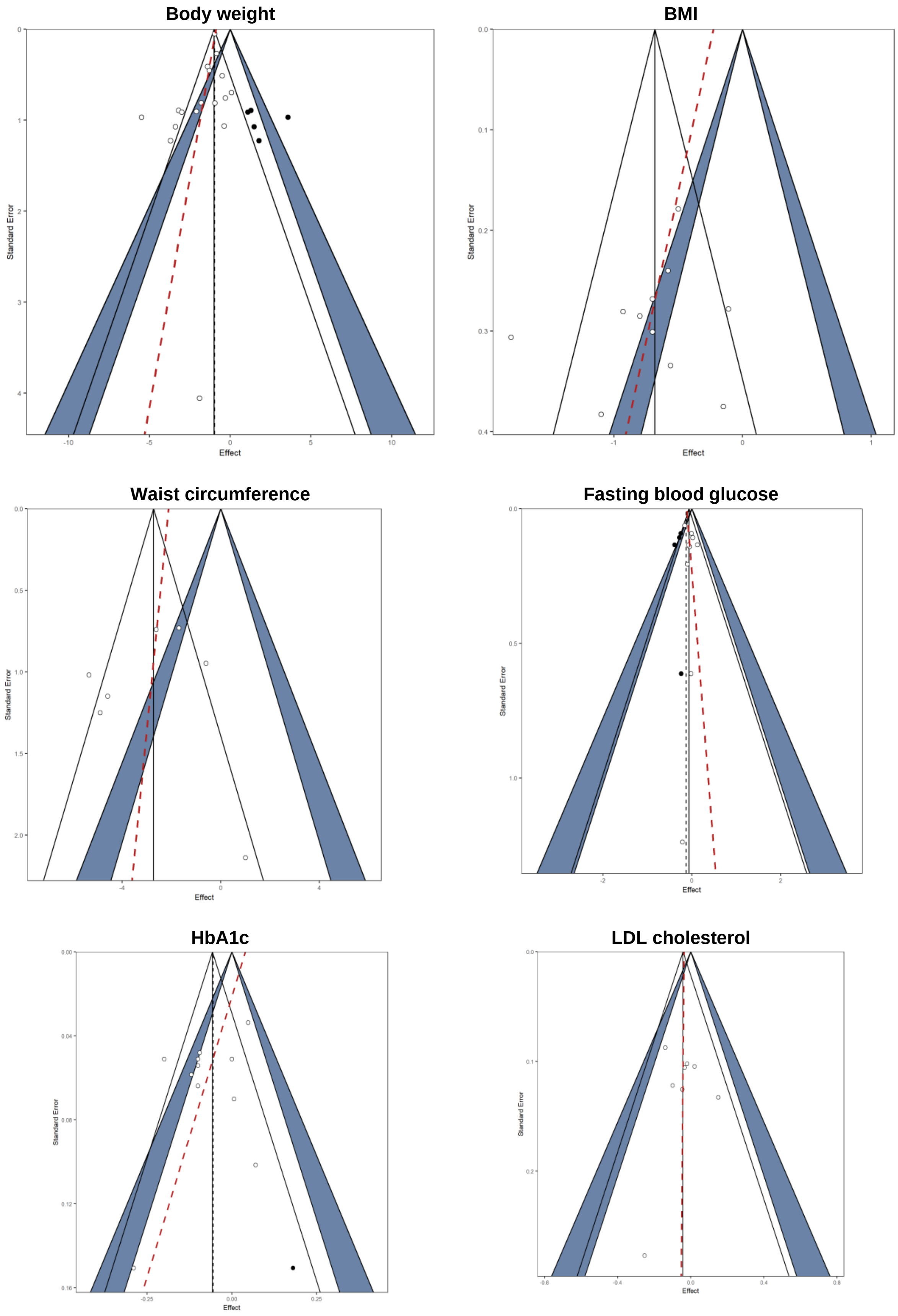

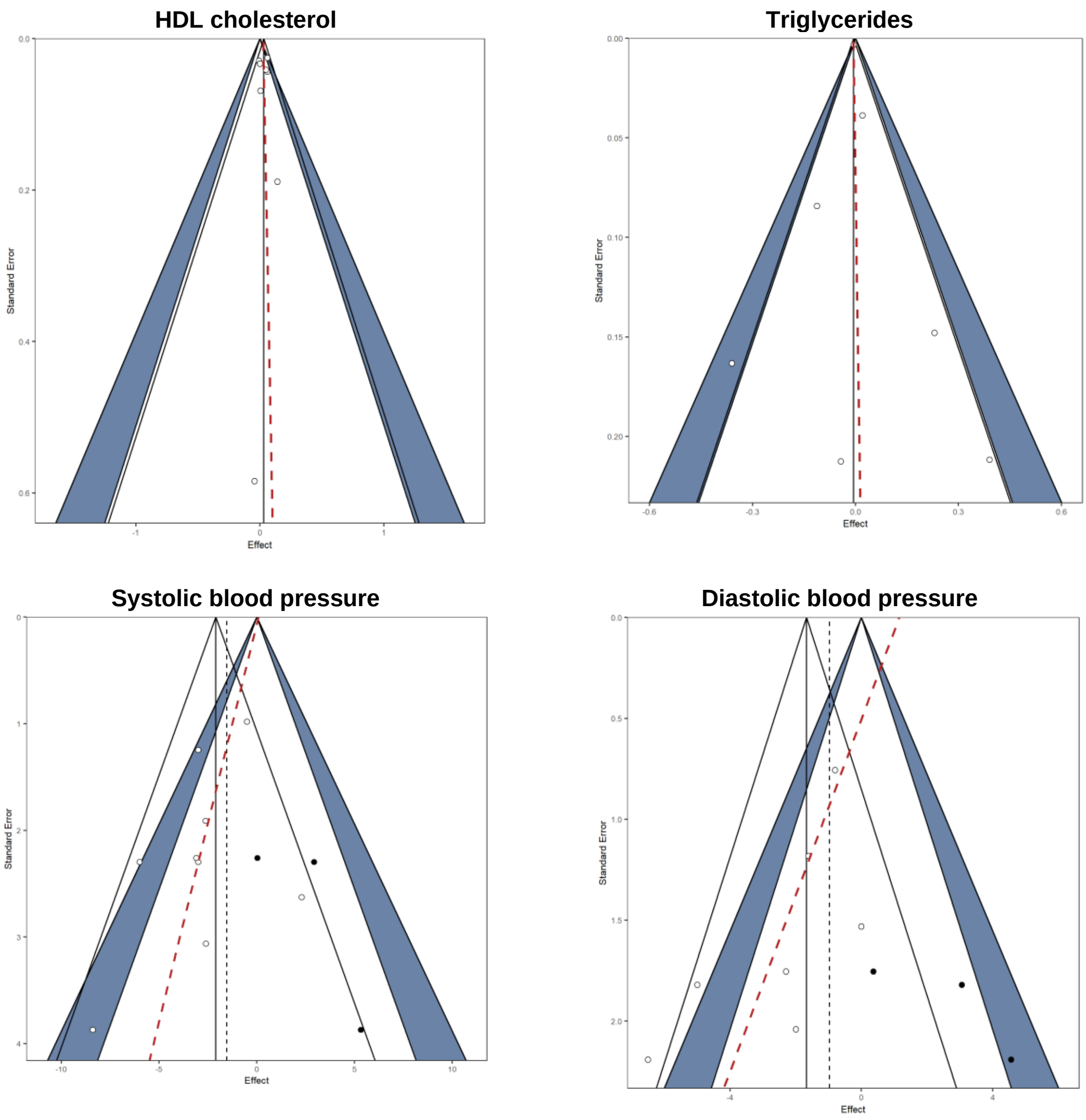

**Supplemental Figure 2.** Funnel plots with trim-and-fill studies included. White dots represent included studies. Black dots represent imputed studies from trim-and-fill. Red broken line is the Egger regression line.
